# Clinical and cost-effectiveness of communication skills e-learning for primary care practitioners on patients’ musculoskeletal pain and enablement: the Talking in Primary care (TIP) cluster-randomised controlled trial

**DOI:** 10.1101/2025.04.09.25325512

**Authors:** Felicity L Bishop, Taeko Becque, Kirsty Garfield, Nadia Cross, Rachel Dewar-Haggart, Emma Teasdale, Amy Herbert, Michelle E Robinson, Matthew J Ridd, Christian Mallen, Lorna Clarson, Jennifer Bostock, Beth Stuart, Leanne Morrison, Sebastien Pollet, Jane Vennik, Helen Atherton, Jeremy Howick, Geraldine M Leydon, Jacqui Nuttall, Nazrul Islam, Paul H Lee, Paul Little, Hazel A Everitt

## Abstract

**Objectives:** To determine the effectiveness and cost-effectiveness of a brief communication skills e-learning package on empathy and optimism for practitioners consulting with adult primary care patients presenting with musculoskeletal (MSK) pain.

**Design:** A cluster randomised controlled trial from 31/10/22 to 28/6/24 using computer-generated block randomisation allocated general practices to intervention or usual care control (1:1) stratified by practice size and deprivation. Patients, trial statisticians, investigators, and researchers involved in collecting outcome data were blinded to allocation. Practitioners were not told which patients were participating in the trial.

**Setting:** General practices in England and Wales (all NHS general practices eligible).

**Participants:** Adults ≥18y consulting a participating practitioner face-to-face, by telephone, or videoconference were recruited into two groups prior to consultation: those consulting about new, recurrent, or ongoing MSK pain and scoring ≥ 4 on an 11-point scale at baseline; and those consulting for any other reason (All-comers). Participating primary care practitioners came from a range of disciplines (e.g., GPs, nurse practitioners, first-contact physiotherapists) and routinely saw patients with MSK pain.

**Interventions:** Intervention arm practitioners received EMPathicO, an evidence-based theoretically-grounded, brief digital e-learning package using behaviour change techniques to enhance communication of clinical empathy and realistic optimism. Control arm practitioners did not receive EMPathicO and consulted patients as usual, access to EMPathicO was provided upon trial completion.

**Main outcomes:** The MSK pain group patient-level dual primary outcomes were pain intensity and patient enablement. The All-comer group patient-level primary outcome was patient enablement. All outcomes were analysed over 6 months using a repeated-measures approach. Cost effectiveness was assessed from UK NHS and societal perspectives including personal expenses and productivity over 6 months.

**Results:** 53 general practices were randomised (25 intervention, 28 control) from which 233 practitioners (115 intervention, 118 control) and 1682 patients were recruited (806 in the MSK pain group (439 intervention, 367 control) and 876 in the All-comers group (490 intervention, 386 control)). Intention to treat analysis found no statistically significant differences between intervention and usual care on primary outcomes. Among the MSK pain group, pain intensity adjusted mean difference was 0.06 (97.5% CI -0.19 to 0.31) and patient enablement adjusted mean difference was 0.17 (97.5% CI -0.05 to 0.40). Among the All-comers group, patient enablement adjusted mean difference was -0.12 (95% CI -0.32 to 0.07). We found no evidence of harm associated with the intervention. From a UK NHS perspective, the probability of cost-effectiveness at a willingness to pay threshold of £20,000 per quality-adjusted life year was 97% for the MSK pain group and 64% for the All-comers group. Compared to control, intervention practitioners had significantly higher self-efficacy for communicating empathy and optimism at 8 weeks (empathy adjusted mean difference was 0.78 (95% CI 0.45 to 1.10), optimism adjusted mean difference was 0.98 (95% CI 0.59 to 1.37)) and 34 weeks post-intervention (these mean differences were 0.63 (95% CI 0.32 to 0.93) and 0.75 (95% CI 0.39 to 1.10), respectively).

**Conclusions:** Brief e-learning for primary care practitioners significantly increased practitioner self-efficacy for a sustained period, is probably cost-effective, and is safe for patients but did not improve pain intensity, patient enablement or other health outcomes. EMPathicO could be rapidly and widely disseminated to support practitioners delivering primary care consultations.

**Registration:** ISRCTN18010240 registered 15 September 2022. Funding: NIHR School for Primary Care Research grant 563.

**Summary:** *What is already known:*

- Empathy education is provided in medical schools and health professional training, yet patients still report limited clinical empathy and related dissatisfaction with their clinical interactions.
- Experimental research suggests realistic optimism has beneficial effects on patient outcomes including pain but how this translates in clinical settings is unknown.
- Existing clinical empathy training is often too lengthy for busy primary care clinicians and cost effectiveness assessments are lacking.

*What this study adds:*

- This large, robust, trial undertaken in UK primary care suggests very brief e-learning for primary care practitioners in clinical empathy and realistic optimism is safe, is probably cost-effective, and significantly enhances practitioner self-efficacy over a sustained period but found no effect on patient outcomes.
- The benefits for practitioners were found across the multidisciplinary primary care team of clinicians, including experienced GPs, GP registrars, physiotherapists and practice nurses from a broad range of GP practices.
- The digital nature of the e-learning package would enable rapid widespread dissemination to clinicians at low cost.

## INTRODUCTION

Successful consultations depend on effective communication. Clear and effective verbal and non-verbal communication is facilitated by clinical empathy – where the practitioner puts themselves in the patient’s position, acknowledges the patient’s feelings, concerns and expectations, and behaves in a way that communicates understanding.^1-3^ Training practitioners in clinical empathy has been shown to increase patient satisfaction,^4^ improve patient symptoms^5^, and foster practitioner personal growth and professional development^6^, and may reduce practitioner burnout.^7-9^ Low levels of clinical empathy may contribute to adverse outcomes including patient and clinician distress, unnecessary or unwanted treatment^10 11^, complaints,^12^ and litigation^13^. Despite decades of research and embedding empathy education in medical schools and health professional training,^14-19^ patients still describe instances of limited empathy in clinical practice which negatively impact their experience.^20 21^

Conveying a positive message where appropriate is measured as part of leading empathy tools^22^ but is rarely emphasised in empathy research or in clinical communication training^23^. Engendering positive expectations of treatment benefit in non-clinical experimental settings can produce measurable neurobiological effects including on the opioid system and decreased reported pain and other symptoms.^24 25^ Whether this could translate into more complex clinical settings warrants further exploration.^26-28^ Communicating clinically realistic optimism involves a practitioner expressing to a patient confidence that a suggested plan of action, such as a prescribed medication or a lifestyle change, will likely lead to the desired patient outcomes, such as less pain or better function;^5 23^ this is seen as novel by primary care practitioners who are keen to learn more.^23^

Existing training in clinical empathy is too lengthy for busy primary care clinicians (training durations averaged 10 hours in a recent review);^29^ targets individual professions rather than the broader, multidisciplinary primary care workforce; is typically delivered face to face; and does not emphasise realistic optimism.^30^ Important questions remain about short and long term effects, e-learning for delivery, and the cost-effectiveness of empathy training.^31^ Larger higher quality clinical trials with public and patient involvement, health economics and nested qualitative work are needed.^3^

EMPathicO is a rigorously developed evidence-based theoretically-grounded brief digital e-learning package in clinical empathy and realistic optimism, for practitioners routinely seeing patients in primary medical care, including GPs, nurse practitioners and first-contact physiotherapists.^20 23 30 32–36^ This trial aimed to address many of the limitations of previous clinical empathy training studies by assessing EMPathicO’s clinical and cost effectiveness in the pressurised clinical UK primary care setting.

## METHODS

### Design

A two-arm cluster randomised controlled trial allocated general practices to intervention or usual care control (1:1). This reduced the cross-contamination risk of individual practitioner or patient level randomisation. Usual care control enabled pragmatic assessment of benefits and costs of adding the EMPathicO e-learning package to usual care. All eligible primary care practitioners in intervention practices were encouraged to complete EMPathicO. Primary care practitioners in control practices consulted as usual and could access the EMPathicO e-learning package after the trial. Primary and secondary outcomes were measured at the level of individual patients. Additional secondary outcomes were measured at the level of individual practitioners. During the trial the sample size calculation was updated to reflect average cluster size, as reported in the published protocol.^37^

Two cohorts of patients were recruited simultaneously: those presenting with musculoskeletal (MSK) pain and all adult consulters (All-comers). MSK pain was chosen due to the high prevalence and burden of MSK pain for patients and health services^38^ and previous evidence of empathy and patient expectations effects on pain^5 24^. All-comers were also included to assess broader potential benefits of practitioners adopting EMPathicO communication skills.

### Participants

#### GP Practices

NHS general practices in England and Wales were eligible. Practices involved in intervention development/feasibility work (18 from Wessex, 5 from West Midlands) and practices where clinical members of the Trial Management Group/Trial Steering Committee see patients (n=5) were ineligible. A diverse sample of practices were sought with local National Institute for Health and Care Research (NIHR) Clinical Research Network (CRN) support, to include large and small, urban and rural practices and practices in areas of higher deprivation and greater ethnic diversity. We required informed consent from at least two eligible practitioners before enrolling a practice in the trial.

#### Practitioners

Eligible practitioners - from any clinical discipline working in a participating general practice and seeing patients with musculoskeletal pain - were invited into the trial by the general practice’s principal investigator. The trial team provided participant information sheets for practitioners and obtained their written informed consent via Qualtrics prior to randomisation. Practitioners unwilling to undertake the intervention/trial procedures were ineligible.

#### Patients

Eligible patients for both MSK pain and All-comers groups were adults (18+); consulting face-to-face, by telephone, or videoconference; able to give informed consent. For the MSK pain group, patients also needed to be consulting a participating practitioner about new, recurrent, or ongoing musculoskeletal pain (e.g. back, hip, upper/lower extremity, neck pain - consistent with ICD-11’s diseases of the musculoskeletal system^39^) and reporting average pain over the last week of 4 or more on a numerical rating scale at baseline (0 = no pain; 10 = pain as bad as you can imagine).^40^ The All-comers group included all other patients consulting a participating practitioner.

Patients were ineligible if they consulted <u>solely</u> in written forms (e.g., e-consult/email); had malignant pain; were unable to consent or complete questionnaires (e.g., severe mental illness or distress, terminal illness); were already enrolled in the trial (i.e., from a previous consultation); were aged <18.

Administrative practice staff invited consecutive patients with prebooked or same-day appointments with a participating practitioner (except patients unable to consent or complete questionnaires). The consultation that prompted the study invitation is termed the ‘index’ consultation. Patients received a brief invitation and link to the Qualtrics-hosted patient-facing study website up to 1 week before their index consultation, via SMS text, email, or post. The website presented the patient information sheet in languages requested by practices (Arabic, Czech, Romanian, Slovak, Somali, Tigrinya, Traditional Chinese, Urdu, Welsh), online consent, screening and baseline questionnaires. Trial staff were available to respond to queries. Participating practitioners were not involved in consenting participants or informed on which patients were participating.

Practices followed their usual procedures for contacting patients with no or limited English, including sending the invitation in the patient’s own language. Professional on-demand interpreters were made available by the trial team (but were not requested by any patients) and informal interpreters were also permitted, to increase research inclusion and remove language barriers to participation.

### Interventions

#### EMPathicO e-Learning Package

EMPathicO aims to help practitioners enhance their communication of clinical empathy and realistic optimism and is consistent with major consultation models including ‘ICE’ (Ideas, Concerns and Expectations).^41^ The brief interactive e-learning modules can be completed separately or together in less than one hour.^42^ Three core modules cover clinical empathy, realistic optimism, tailoring empathy and optimism for patients with osteoarthritis (a common cause of musculoskeletal pain, showing clear demonstration of communication skills in a particular context).^43^ Subsequent activities guide practitioners to evaluate their own consultations, set goals, and review their goals. EMPathicO was designed to target users’ motivation (reflective, autonomic), capability (physical, psychological), and opportunity (environmental), through intervention functions of persuasion, incentivisation, enablement, education, training, modelling, and environment restructuring. Multiple behaviour change techniques were used to achieve these functions, including demonstration, information provision, goal-setting, action planning, and instruction. EMPathicO was deployed on the LifeGuide+ platform for developing and evaluating digital interventions.^44^ The development of EMPathicO was described here;^32^ the version used in this trial is described as per TIDieR (see Supplementary Material).

#### Control: Usual Care

Practitioners in practices randomised to the usual care control arm were instructed to “continue to treat patients as usual and not undertake any training in communication skills.” They were given access to the EMPathicO e-learning package after all patient recruitment and follow-up was completed.

#### Concomitant Interventions

All practitioners were discouraged from undertaking additional communication skills training during the study and to self-report any that did occur. Fewer than 15% of practitioners reported doing additional communication skills training (intervention group: 7/64 (10.9%); control group: 11/74 (14.9%)).

#### Outcomes

Patient-reported measures were completed online on Qualtrics (Qualtrics, Provo, UT); to support inclusive access patients could request paper versions. Patients completed questionnaires at pre-consultation baseline (up to 7 days before index consultation) and 7 days, 1 month, 3 months, and 6 months after index consultation. Paper versions were completed by 3 patients at baseline, 11 at 7d, 12 at 1m, 10 at 3m, and 13 at 6m. £10 vouchers were sent at 1-month and 6-months, unconditional on response, to incentivise completion. Automated follow-up emails were sent to non-responders at 3 and 7 days. From 10 days, researchers personally contacted persistent non-withdrawn non-responders, offering to resend questionnaires or complete primary outcomes by telephone. While links to questionnaires were resent for many participants, only one baseline questionnaire was completed by telephone; no other questionnaires were completed by telephone. Practitioner-reported measures were completed at baseline, 2 weeks, 8 weeks, and 34 weeks, online on LifeGuide+ (intervention group-only measures) and Qualtrics (all other practitioner reported outcomes). Non-responders were sent at least two reminders: at 2w, two automated reminders were timed contingent on intervention progress; at 8w and 34w, automated reminders were sent at 3 and 7 days, personalised reminders at 14 and 21 days. At 34w, principal investigators at each practice were also contacted to encourage questionnaire completion.

#### Primary outcomes

For the MSK pain group, the dual primary outcomes were patient reported pain intensity and enablement, each analysed over 6 months using a repeated measures approach. Pain intensity, the severity of pain sensation, was operationalised as average pain in the last week measured using the Brief Pain Inventory (BPI).^40^ Patient enablement was measured on the 6-item Patient Enablement Index (PEI)^45^ with 7-point response options,^37 46-48^ capturing patients’ feelings after a consultation, of confidence and empowerment to cope with their symptoms, to keep healthy and to help themselves.

For the All-comers group, patient enablement was the single primary outcome. Pain intensity was measured as a secondary outcome if pain was present.

Pain intensity was measured at baseline and all follow-ups, and patient enablement was measured at all follow-ups.

#### Secondary outcomes

Patient-reported secondary outcomes were: pain intensity at all time-points (BPI 4-item pain intensity subscale);^40^ perceived symptom severity measured at all time-points and symptom change measured at all follow-ups (7-point^49^ Patient Global Impression of Symptom Severity and Patient Global Impression of Change);^50^ patient satisfaction with the consultation at <7d follow-up (21-item Medical Interview Satisfaction Scale^51^ for UK primary care);^52^ and pain interference at 1m and 6m follow-up (BPI 7-item pain interference subscale).^40^

#### Health economics outcomes

Patient health-related quality of life (5-item EQ-5D-5L, EQ-VAS)^53^ and capability wellbeing (5-item ICECAP-A)^54 55^ were measured at baseline, 1m and 6m. Resource-use data was collected at 3m and 6m (ModRUM^56^ with bespoke questions for personal expenses; Work Productivity and Activity Impairment Questionnaire: General Health)^57^. Practitioner time spent on the EMPathicO e-learning package was captured by the host platform.

#### Process measures

Patient-reported process measures at <7d follow-up were: perceptions of practitioner clinical empathy (measured on the 10-item CARE);^22^ patient perceptions of practitioner response expectancies (bespoke item developed for this trial); patient treatment outcome expectancies (15-item 6-subscale, Treatment Expectation Questionnaire (TEX-Q);^58^ patient anxiety and depression (7-item subscales of the Hospital Anxiety and Depression Scale (HADS));^59 60^ continuity of care (9-item Patient-Doctor Depth of Relationship PDDR Scale^61^ modified for non-doctor practitioners). Patients reported at baseline their age, gender, ethnicity, and postcode (for calculating index of multiple deprivation, IMD) and at <7d follow-up their reason(s) for consulting (coded using the ICPC-2), comorbidities and index consultation modality.

All practitioners reported self-efficacy for conveying empathy and optimism in consultations at pre-randomisation baseline, 8w and 34w post-randomisation (using bespoke scales previously tested)^42^. Data collected from intervention arm participants only are reported elsewhere^62^ and included: outcome expectancy and intentions for conveying empathy and optimism in consultations (at 2w, 8w, and 34w post-randomisation, using bespoke scales previously tested);^42^ self-reported change in communicating empathy and optimism in consultations (8w and 34w), and extent and patterns of use of the EMPathicO e-learning package (captured by LifeGuide+). Practitioners reported at baseline their age, gender, ethnicity, years qualified, and profession. Internal consistency of all bespoke scales was acceptable (Cronbach’s alphas ranged from .81 to .96).

Practice-level characteristics were collected from practices and supplemented with data from national general practice profiles (National General Practice Profiles - Data – OHID, phe.org.uk): list size, deprivation score, staffing.

Non-prespecified outcomes were explored in the nested qualitative study.^63^

#### Sample Size

For the MSK pain group, the minimum clinically important difference in the pain primary outcome is approximately one point.^64^ Assuming a standard deviation of 3.3, this difference is equivalent to a standardised effect size of 0.3. To achieve 90% power, alpha 0.025 to allow for two primary outcomes, and a correlation between the 4 repeated measures of 0.7, a sample size of 214 per group was required for an individually-randomised trial. In our clustered RCT, we assumed a conservative ICC of 0.03^65^, assumed 14 patients per practice yielding a design effect of 1.39, and allowed for 20% loss to follow up, meaning (214*2*1.39)/0.8=744 participants are needed to be recruited from 53 practices. Recruiting 744 All-comers would give more than 90% power (based on alpha 0.05 and ICC 0.03) to detect a standardised effect size of 0.3 in the enablement primary outcome, equivalent to a difference of 0.36 points (assuming SD=1.2^66^).

#### Randomisation and Blinding

The computer-generated randomisation sequence used blocks (block sizes 4 and 6) stratified by practice-level deprivation (IMD 1-5 / IMD 6-10) and practice size (list size>7900 / <7900; 7900 = median practice list size in England). The allocation sequence was programmed by an independent programmer in LifeGuide+ and was not visible to users. The trial manager (or their delegate) enrolled clusters (i.e., practices) and obtained electronic written consent from practitioners prior to randomisation. The trial manager then entered practices into the randomisation function on LifeGuide+ and communicated the allocation to practices.

Patients, trial statisticians, investigators, and researchers collecting patient outcomes were blinded to allocation until after analysis of primary outcomes. The research team did not tell practitioners which patients were participating and patients were instructed: “please do not discuss your participation in the study with your GP, nurse, physiotherapist, or any other primary care practitioner”. Patients were not told in advance that some practitioners had received communication skills training as part of this study. This was ethically approved as appropriate for a cluster-randomised trial of a low-risk communication-skills training intervention within the broad scope of usual practice.

### Statistical Methods

MSK and All-comers groups were analysed separately. The primary analyses for the BPI and PEI scores used a generalized linear mixed model (GLMM) framework with observations at 3 days, 1-. 3-, and 6-months (level 1) nested in patients (level 2) and patients nested in practices (level 3). This model used all the available data, implicitly assuming that missing outcome scores are missing at random given the observed data. The model adjusted for baseline BPI and stratification variables (practice size and practice deprivation). An unstructured covariance matrix was used to avoid imposing any constraints on the covariance. For secondary outcomes, the analyses used a similar generalised linear mixed modelling approach, controlling for baseline values (for continuous outcomes), and stratification variables. Pre-planned exploratory subgroup analyses were undertaken by age, gender, deprivation, baseline pain, consultation reason, consultation modality and practitioner role. Intention to treat analysis (as randomised) was undertaken regardless of any practice-level non-adherence to the intervention.

Practitioner-reported outcomes were analysed at 8 and 34 weeks using linear mixed models, with practitioners nested within practices, and adjusting for baseline values and stratification factors. Internal consistency of the measures was assessed using Cronbach’s alpha.

Analyses were conducted in Stata 18. A full and detailed statistical analysis plan was developed prior to final trial analysis, approved by Trial Steering Committee and published on ISRCTN.

### Health Economic Methods

Cost effectiveness was assessed, separately for MSK and All-comers groups, from a primary NHS perspective and secondary societal perspectives including private healthcare, travel costs for healthcare, over-the-counter medications and productivity over 6 months. Utility scores were estimated from EQ-5D-5L scores using the NICE-recommended mapping function.^67-69^ Quality-adjusted life years (QALYs) were estimated by combining utility values, with length of time in each health state, using the area under the curve approach.^53^ ^70–74^ Intervention resources included practitioner time and the LifeGuide platform (including hosting, support and domain name). To estimate intervention costs it was conservatively assumed 25% of clinical staff would undertake training at each practice. NHS resources and sick leave from employment were valued using 2022/23 national unit costs (Supplementary Table 7).^74-76^ Personal expenses are presented as reported. Discounting was not required.

All analyses were conducted in Stata 18, on an intention-to-treat basis. In the primary analysis, missing data was imputed using multiple imputation by chained equations. All analyses were adjusted for stratification factors. Data distribution was handled using generalised linear models, and the cluster design was accounted for using a clustered sandwich estimator. Incremental net monetary benefits (INMBs) were estimated at £20,000 and £30,000 per QALY. In cost-effectiveness analyses, cost per year of full capability equivalent gained was planned to be estimated if EMPathicO was more costly and more effective. Uncertainty was explored via cost-effectiveness acceptability curves and sensitivity analyses, which included a complete case analysis and an analysis from an NHS primary care perspective.

### Data and Code Sharing

Data and code are available on request. Requests for deidentified participant data may be submitted to the University of Southampton data repository, quoting doi: [to be added on acceptance]. Requests would be subject to review by a subgroup of the trial team. Access to anonymised data may be granted following this review, subject to conditions including ethical approval, qualifications, and aims consistent with the original purpose of the study. All data-sharing activities would require a data-sharing agreement. Code will be shared via GitHub repository.

### Patient and Public Involvement

To ensure our work engages and is relevant to patients, we have worked with patients and members of the public throughout developing and trialling EMPathicO. Pre-trial, our patient advisors influenced the choice of dual primary outcomes for the musculoskeletal pain group. During the trial, our Patient Advisory Group was led by JB (patient, co-author, and member of trial management group) and comprised six patient and public contributors of varying ages, ethnic backgrounds (three from minority ethnic groups, three from White backgrounds), gender (three female, three male), and geographical locations within England. One member is neurodivergent, and all have lived experience of musculoskeletal pain as patients or carers and had positive and negative experiences of communication in primary care. The group met virtually bimonthly and contributed to specific activities including refining patient-facing documents and procedures, piloting qualitative interviews, interpreting data and dissemination activities.

## RESULTS

### Participant Recruitment and Flow

In total, 53 practices and 233 practitioners participated. Figure 1 shows practice and individual practitioner flow. Practices were randomised between 31/10/22 and 17/10/23. No practices withdrew after randomisation. One hundred and eleven practitioners (96.5%) of the one hundred and fifteen allocated to the intervention worked through all three content modules of the EMPathicO e-learning package (the remainder withdrew; see companion paper for more details on intervention usage). Eighteen practitioners withdrew after randomisation: 1 no longer had capacity for research and 17 were no longer seeing patients at the practice, having either left the practice, taken sick leave, taken maternity leave, or died.

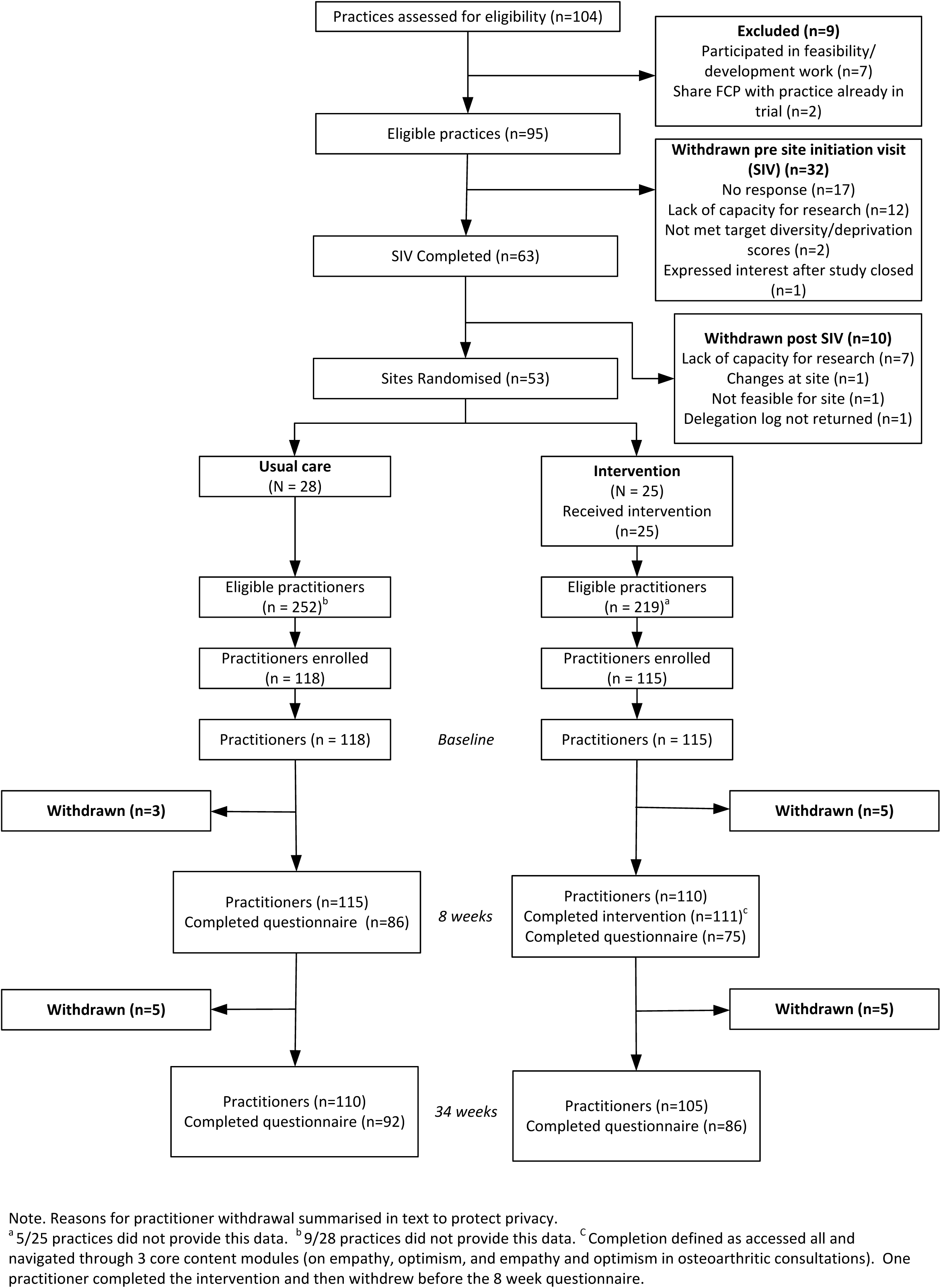

In total, 1682 patients participated (806 in the MSK pain group; 876 in the usual care group). Figure 2 shows individual patient flow. Patients were recruited between 15/11/22 and 18/12/23 and were followed up for 6 months. Patient follow-up was completed on 28/6/24. Recruitment ceased on reaching the required sample size in the MSK pain group. Over 6 months, in the MSK pain group 9/367 (2.5%) withdrew from the intervention arm and 5/439 (1.1%) withdrew from the usual care arm. In the All-comers group, 7/490 (1.4%) withdrew from intervention and 6/368 (1.6%) withdrew from usual care.

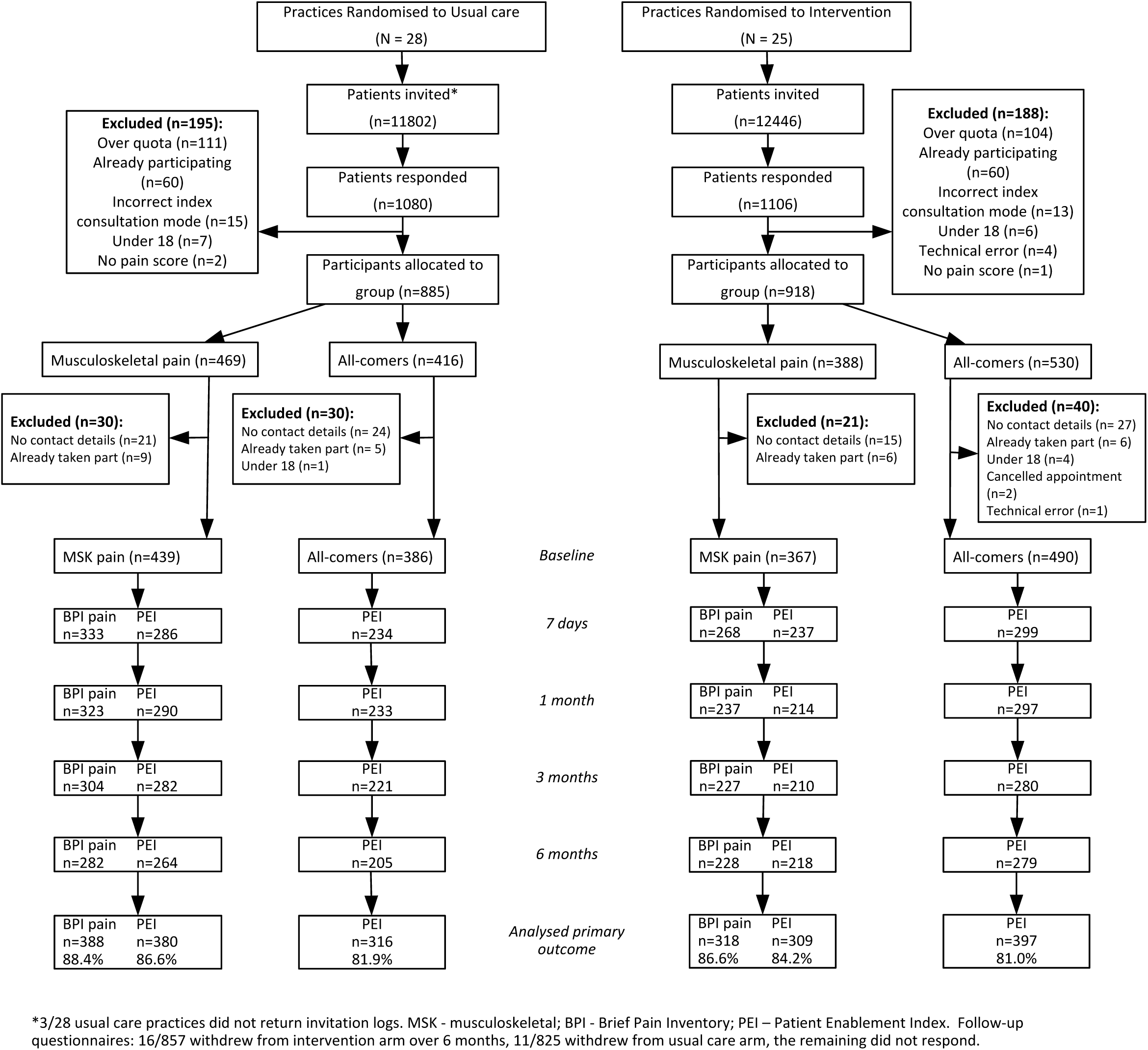

### Baseline Data

Practice size and deprivation score and most practitioner characteristics were well balanced between randomised arms (Table 1). A range of primary care practitioners took part including GPs, first contact practitioners, and nurse practitioners. There was a slightly higher proportion of female practitioners in the intervention arm compared to the control arm. On average, 4 practitioners and 38% of eligible practitioners took part from each practice.

**Table 1.** Practice and Practitioner Baseline Characteristics.

|  | Usual care<br>n (%) | Intervention<br>n (%) | Overall<br>n (%) |
| --- | --- | --- | --- |
| **Practice Characteristics** |  |  |  |
| Practices, n | 28 | 25 | 53 |
| Practice size |  |  |  |
| Large (list size >=7900) | 17 (60.7) | 15 (60.0) | 32 (60.4) |
| Small (list size <7900) | 11 (39.3) | 10 (40.0) | 21 (39.6) |
| Practice deprivation score |  |  |  |
| Higher (IMD deciles 1-5) | 11 (39.3) | 9 (36.0) | 20 (37.7) |
| Lower (IMD deciles 6-10) | 17 (60.7) | 16 (64.0) | 33 (62.3) |
| Practitioner participants |  |  |  |
| Number per practice, median (IQR) | 4 (2, 5.5) | 4 (3, 5) | 4 (3, 5) |
| Proportion of eligible per practice, median (IQR) | 0.35 (0.23, 0.67) | 0.40 (0.28, 0.73) | 0.38 (0.23, 0.67) |
| **Practitioner Characteristics** |  |  |  |
| Practitioners, n | 118 | 115 | 233 |
| Age, mean (SD) | 42.2 (9.2) | 43.4 (8.8) | 42.8 (9.0) |
| Missing, n | 2 | 2 | 4 |
|  | Usual care<br>n(%) | Intervention<br>n(%) | Overall<br>n(%) |
| Gender |  |  |  |
| Male | 59 (50.4) | 42 (36.8) | 101 (43.7) |
| Female | 58 (49.6) | 72 (63.2) | 130 (56.3) |
| Missing, n | 1 | 1 | 2 |
| Ethnicity |  |  |  |
| White | 94 (80.3) | 92 (80.7) | 186 (80.5) |
| Mixed / Multiple | 6 (5.1) | 5 (4.4) | 11 (4.8) |
| Asian / Asian British | 14 (12.0) | 13 (11.4) | 27 (11.7) |
| Black/ African/ Caribbean | 1 (0.9) | 3 (2.6) | 4 (1.7) |
| Other | 2 (1.7) | 1 (0.9) | 3 (1.3) |
| Missing, n | 1 | 1 | 2 |
| Years in role, median (IQR) | 3 (2-9) | 5 (1-10) | 4 (1-10) |
| Missing, n | 2 | 1 | 3 |
| Years qualified, median (IQR) | 10 (4-17) | 12 (3-19) | 10 (4-18) |
| Missing, n | 4 | 4 | 8 |
| Job role |  |  |  |
| GP partner | 47 (40.2) | 49 (43.0) | 96 (41.6) |
| Salaried GP | 15 (12.8) | 18 (15.8) | 33 (14.3) |
| GP trainee/<br>retainer/registrar | 9 (7.7) | 6 (5.3) | 15 (6.5) |
| Nurse practitioner/Nurse | 14 (12.0) | 11 (9.6) | 25 (10.8) |
| Physiotherapist | 21 (18.0) | 16 (14.0) | 37 (16.0) |
| First Contact Practitioner | 2 (1.7) | 2 (1.8) | 4 (1.7) |
| Paramedic | 4 (3.4) | 6 (5.3) | 10 (4.3) |
| Pharmacist | 2 (1.7) | 2 (1.8) | 4 (1.7) |
| Physician associate | 1 (0.9) | 3 (2.6) | 4 (1.7) |
| Other | 2 (1.7) | 1 (0.9) | 3 (1.3) |
| Missing, n | 1 | 1 | 2 |

**Table 2.** Baseline Characteristics of Patients in the Musculoskeletal Pain Group.

|  |  |  |  |
| --- | --- | --- | --- |
| Participants, n | 439 | 367 | 806 |
| Age |  |  |  |
| Median (IQR) | 62 (53-70) | 61 (51-70) | 62 (52-70) |
| Mean (SD) | 60.1 (13.9) | 59.1 (14.5) | 59.7 (14.2) |
| Range | 18-89 | 21-92 | 18-92 |
| Gender |  |  |  |
| Male | 125 (28.5) | 104 (28.3) | 229 (28.4) |
| Female | 313 (71.3) | 261 (71.1) | 574 (71.2) |
| Prefer not to say | 0 (0.0) | 1 (0.3) | 1 (0.1) |
| Other | 1 (0.2) | 1 (0.3) | 2 (0.2) |
| Ethnicity |  |  |  |
| White | 427 (97.3) | 339 (92.4) | 766 (95.0) |
| Mixed / Multiple ethnic groups | 4 (0.9) | 7 (1.9) | 11 (1.4) |
| Asian / Asian British | 5 (1.1) | 5 (1.4) | 10 (1.2) |
| Black/ African / Caribbean / Black British | 1 (0.2) | 9 (2.5) | 10 (1.2) |
| Other Ethnic Group | 2 (0.5) | 7 (1.9) | 9 (1.1) |
| Educational level |  |  |  |
| No formal educational qualifications | 37 (8.4) | 49 (13.4) | 86 (10.7) |
| GCSEs /O-levels or similar | 143 (32.6) | 122 (33.2) | 265 (32.9) |
| A-levels or similar or ONC / OND | 68 (15.5) | 52 (14.2) | 120 (14.9) |
| HNC / HND Degree | 29 (6.6) | 23 (6.3) | 52 (6.5) |
| Degree | 86 (19.6) | 66 (18.0) | 152 (18.9) |
| Higher degree | 24 (5.5) | 17 (4.6) | 41 (5.1) |
| Postgraduate degree | 40 (9.1) | 22 (6.0) | 62 (7.7) |
| Other | 12 (2.7) | 16 (4.4) | 28 (3.5) |
| Employment status |  |  |  |
| Employed full-time | 107 (24.4) | 90 (24.6) | 197 (24.5) |
| Employed part-time | 72 (16.4) | 52 (14.2) | 124 (15.4) |
| Retired | 190 (43.3) | 138 (37.7) | 328 (40.7) |
| Unemployed | 7 (1.6) | 14 (3.8) | 21 (2.6) |
| Doing unpaid work (eg volunteering) | 3 (0.7) | 9 (2.5) | 12 (1.5) |
| Unable to work | 34 (7.7) | 38 (10.4) | 72 (8.9) |
| Other | 26 (5.9) | 25 (6.8) | 51 (6.3) |
| Missing | 0 | 1 | 1 |
| Index multiple deprivation, median (IQR) | 7 (5-9) | 6 (4-8) | 7 (4-9) |
| Pain intensity last week (BPI), mean (SD) | 6.3 (1.5) | 6.3 (1.6) | 6.3 (1.6) |
| Pain intensity subscale (BPI), mean (SD) | 6.0 (1.6) | 6.0 (1.7) | 6.0 (1.6) |
| Symptom severity (PGIS), mean (SD) | 4.4 (1.1) | 4.5 (1.1) | 4.4 (1.1) |
| Self-reported main site of musculoskeletal pain* |  |  |  |
| Neck pain | 19 (4.3) | 7 (1.9) | 26 (3.2) |
| Back pain | 27 (6.2) | 37 (10.1) | 64 (8.0) |
| Low back pain | 45 (10.3) | 37 (10.1) | 82 (10.2) |
| Muscular chest pain | 3 (0.7) | 2 (0.6) | 5 (0.6) |
| Flank/axilla pain | 3 (0.7) | 1 (0.3) | 4 (0.5) |
| Jaw pain | 1 (0.2) | 0 (0.0) | 1 (0.1) |
| Shoulder pain | 50 (11.4) | 36 (9.9) | 86 (10.7) |
| Arm pain | 6 (1.4) | 10 (2.7) | 16 (2.0) |
| Elbow pain | 6 (1.4) | 10 (2.7) | 16 (2.0) |
| Wrist pain | 14 (3.2) | 13 (3.6) | 27 (3.4) |
| Hand/finger pain | 20 (4.6) | 15 (4.1) | 35 (4.4) |
| Hip pain | 43 (9.8) | 41 (11.2) | 84 (10.5) |
| Leg/thigh pain | 21 (4.8) | 20 (5.5) | 41 (5.1) |
| Knee pain | 89 (20.3) | 69 (18.9) | 158 (19.7) |
| Ankle pain | 17 (3.9) | 8 (2.2) | 25 (3.1) |
| Foot/toe pain | 25 (5.7) | 15 (4.1) | 40 (5.0) |
| Musculoskeletal pain | 44 (10.1) | 37 (10.1) | 81 (10.1) |
| Other muscle/bone | 5 (1.1) | 7 (1.9) | 12 (1.5) |
| Appointment modality |  |  |  |
| In person | 361 (82.2) | 311 (84.7) | 672 (83.4) |
| Telephone | 78 (17.8) | 56 (15.3) | 134 (16.6) |

BPI – Brief Pain Inventory scale 0 (no pain) to 10 (most pain); PGIS - Patient Global Impression of Severity scale 1 (none) to 7 (extremely severe); IQR interquartile range (quartile 1, quartile 3); SD standard deviation.
\*Options based on ICPC-2.

### Primary and Secondary Outcomes

#### Patient outcomes: Musculoskeletal pain group

In the MSK pain group, follow up rates (i.e., at least one post-baseline follow-up completed) for the dual primary outcomes exceeded 80% (pain: 318/367 (86.6%) in intervention arm and 388/439 (88.4%) in usual care; enablement: 309/367 (84.2%) in the intervention arm and 380/439 (86.6%) in usual care). There was no evidence of any difference between intervention and usual care for pain intensity (adjusted mean difference 0.06, 97.5% CI -0.19 to 0.31; ICC 0.034) or patient enablement (adjusted mean difference 0.17, 97.5% CI -0.05 to 0.40, ICC 0.027). Due to the imbalance in numbers of patients across randomised arms, a post-hoc analysis adjusting for age, gender, ethnicity, education, employment, IMD, appointment type was undertaken, but did not change the inferences (adjusted mean difference for pain intensity 0.0005, 97.5% CI -0.25 to 0.25, and enablement 0.18, 97.5% CI -0.03 to 0.40). There were also no statistically significant differences between the intervention and usual care arms for BPI mean pain, BPI interference, Patient Global Impression of Severity, Patient Global Impression of Change, patient satisfaction or the patient-reported process measures (Table 3). Use of pain medications was broadly comparable between the intervention and usual care arms (Supplementary Table 2).

**Table 3.**
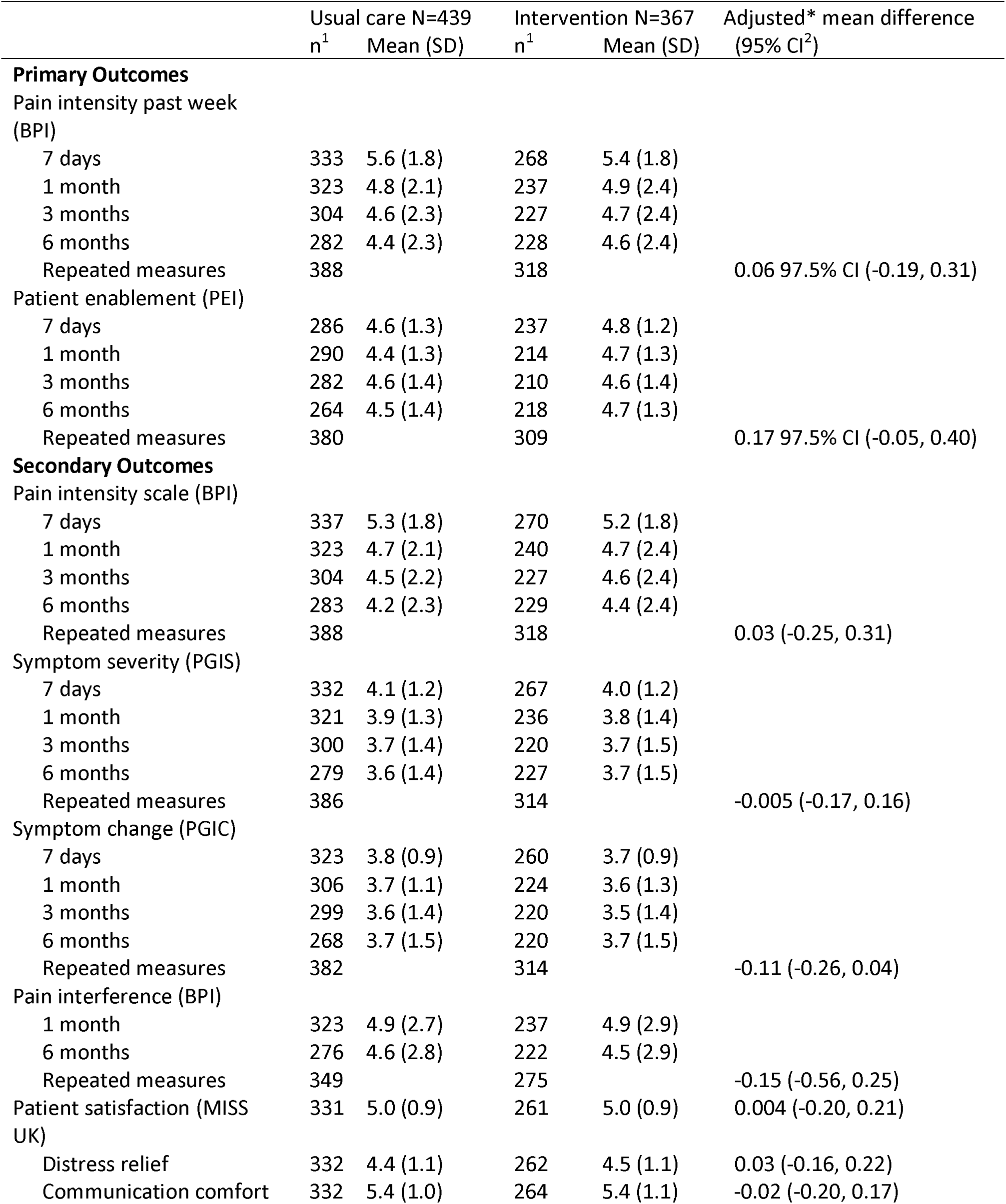
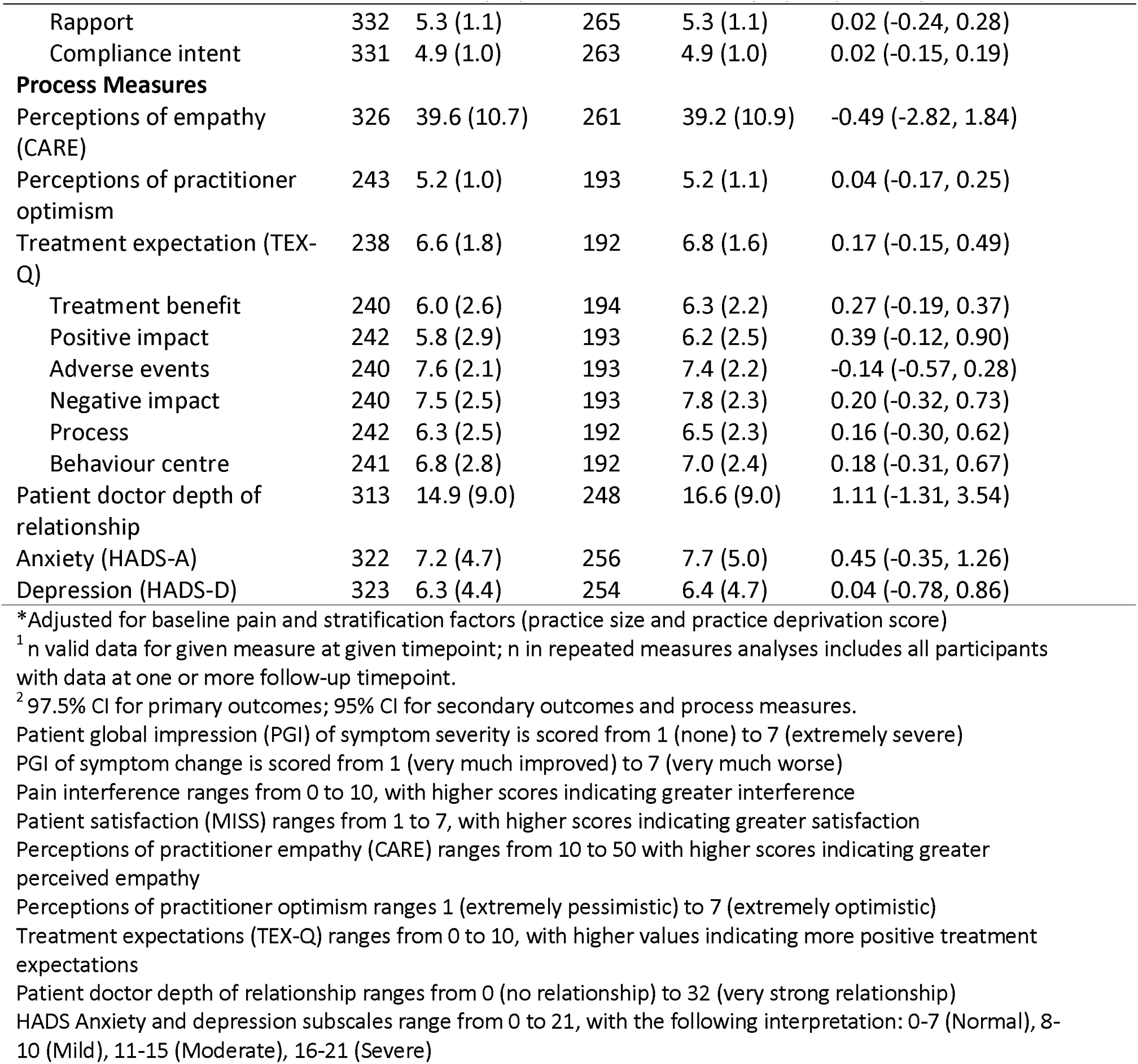
Musculoskeletal group primary and secondary outcomes by intervention allocation.

#### Patient outcomes: All-comers group

In the All-comers group, there was no evidence of any difference in patient enablement between intervention and usual care arms (adjusted mean difference -0.12, 95% CI -0.32 to 0.07). A post-hoc analysis adjusting for age, gender, ethnicity, education, employment, IMD, appointment type did not change the inferences (adjusted mean difference PEI -0.04, 95% CI -0.35 to 0.26). There were also no statistically significant differences between the intervention and usual care arms for Patient Global Impression of Severity, Patient Global Impression of Change, patient satisfaction or the patient-reported process measures (Supplementary Table 3).

#### Practitioner-reported outcomes

Practitioners in the intervention arm had higher self-efficacy for communicating clinical empathy and realistic optimism than practitioners in the control arm, at both 8- and 34-weeks post-randomization (Table 4).

**Table 4.**
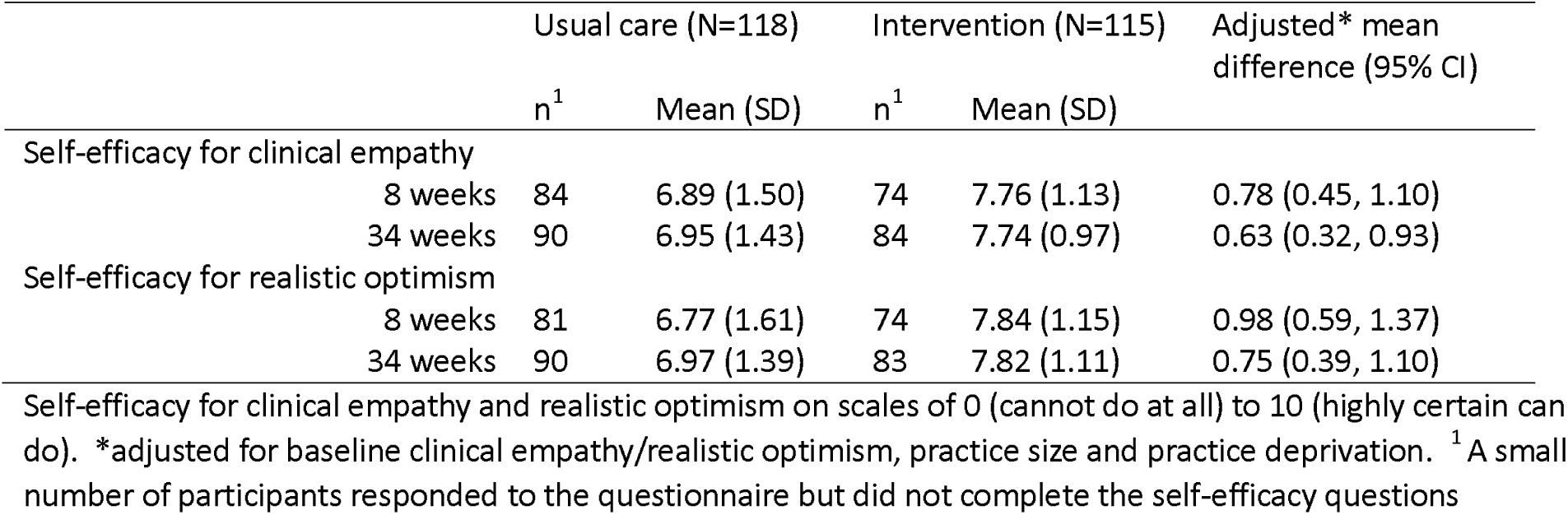
Practitioner reported self-efficacy and attitudes related to clinical empathy and realistic optimism.

#### Subgroup Analyses

In terms of the preplanned subgroup analyses in the MSK pain group, females had less improvement in BPI average pain scores than males, interaction term 0.48, 97.5% CI (0.03, 0.94); and middle aged and older age groups (45-65 and 65+) had a greater improvement in BPI average pain scores than the younger age group (<45 years), adjusted interaction term -0.72 (-1.34, -0.11) (Supplementary Table 4). There were no other statistically significant differences in the subgroups for pain or for enablement (Supplementary Table 5).

### Harms

All serious adverse events were deemed unrelated to the intervention. There was no evidence of any difference in the proportion of those experiencing a serious adverse event: 3.3% intervention vs 3.4% usual care (odds ratio 0.93 95% CI (0.43, 2.03)) for the MSK participants, and 3.5% intervention vs 4.2% usual care (OR 0.84 95% CI (0.42, 1.68)) for the All-comer participants (Supplementary Table 6).

### Health Economic Analyses

Mean practitioner time spent on the EMPathicO e-learning package was 81 minutes. The cost per patient was low (£0.42).

In the MSK group, based on complete case data, mean primary care costs were similar between arms, but mean secondary care costs were somewhat larger in the usual care arm (Table 5). Adjusted total health care costs were lower in the intervention arm, but confidence intervals were wide, indicating no strong evidence of a difference (adjusted incremental difference -£104; 95% CI -£393 to £186). Adjusted QALYS were higher in the intervention arm, but confidence intervals crossed zero indicating no definitive evidence of a difference (adjusted incremental difference 0.012; 95% CI - 0.002 to 0.025). From an NHS perspective, the INMB at a willingness-to-pay threshold of £20,000 was £423 (95% CI -£9 to £854), with a 97% probability of being cost effective. Uncertainty is presented in the CEAC (Figure 3). Mean resource use and costs are presented in Supplementary Table 7 and health economic outcomes are presented in Supplementary Table 8. Mean societal costs were similar between arms (Supplementary Table 9).

**Table 5.**
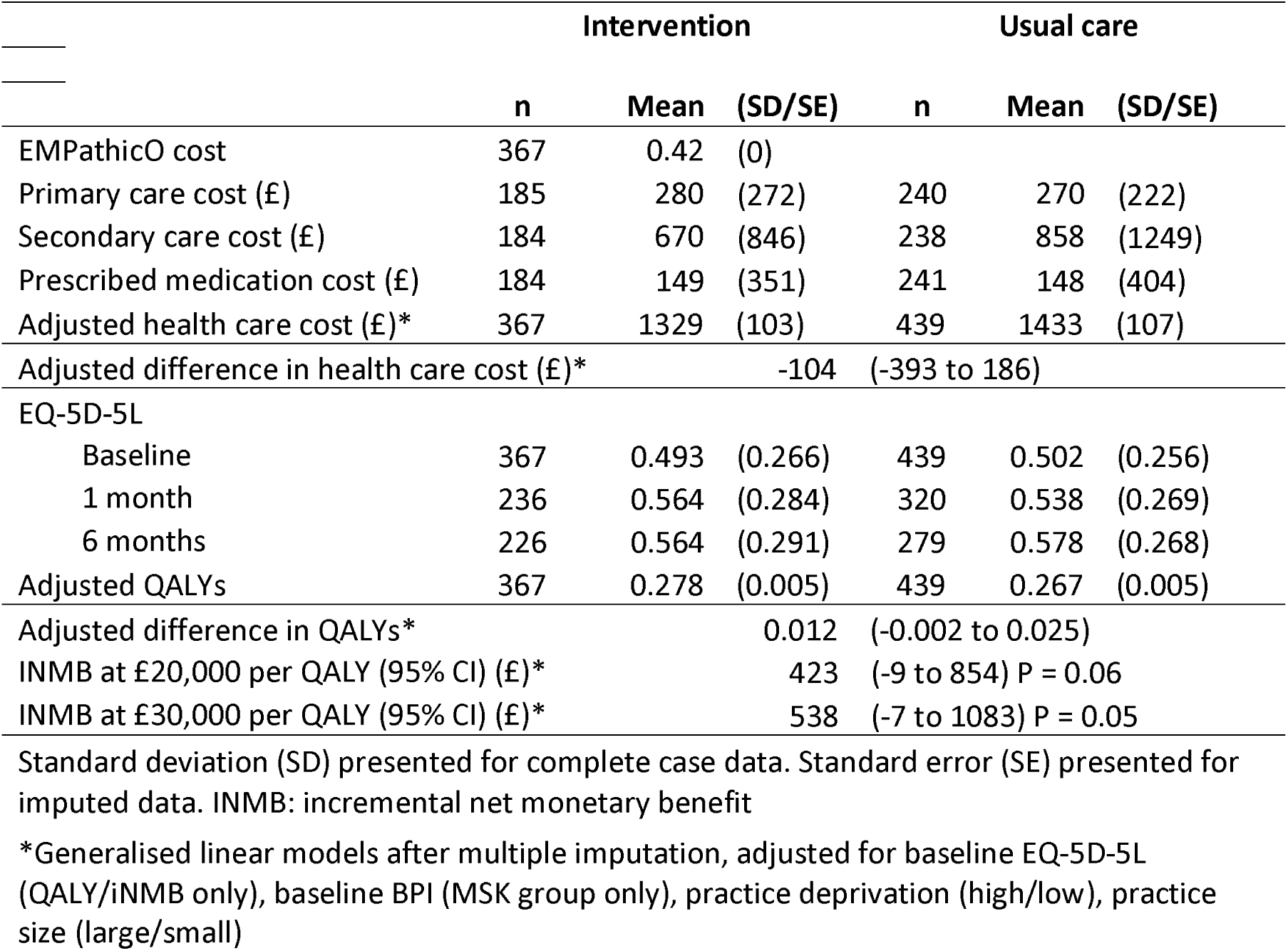
Incremental costs, QALYs and net benefit from an NHS perspective, by intervention allocation for musculoskeletal pain group.

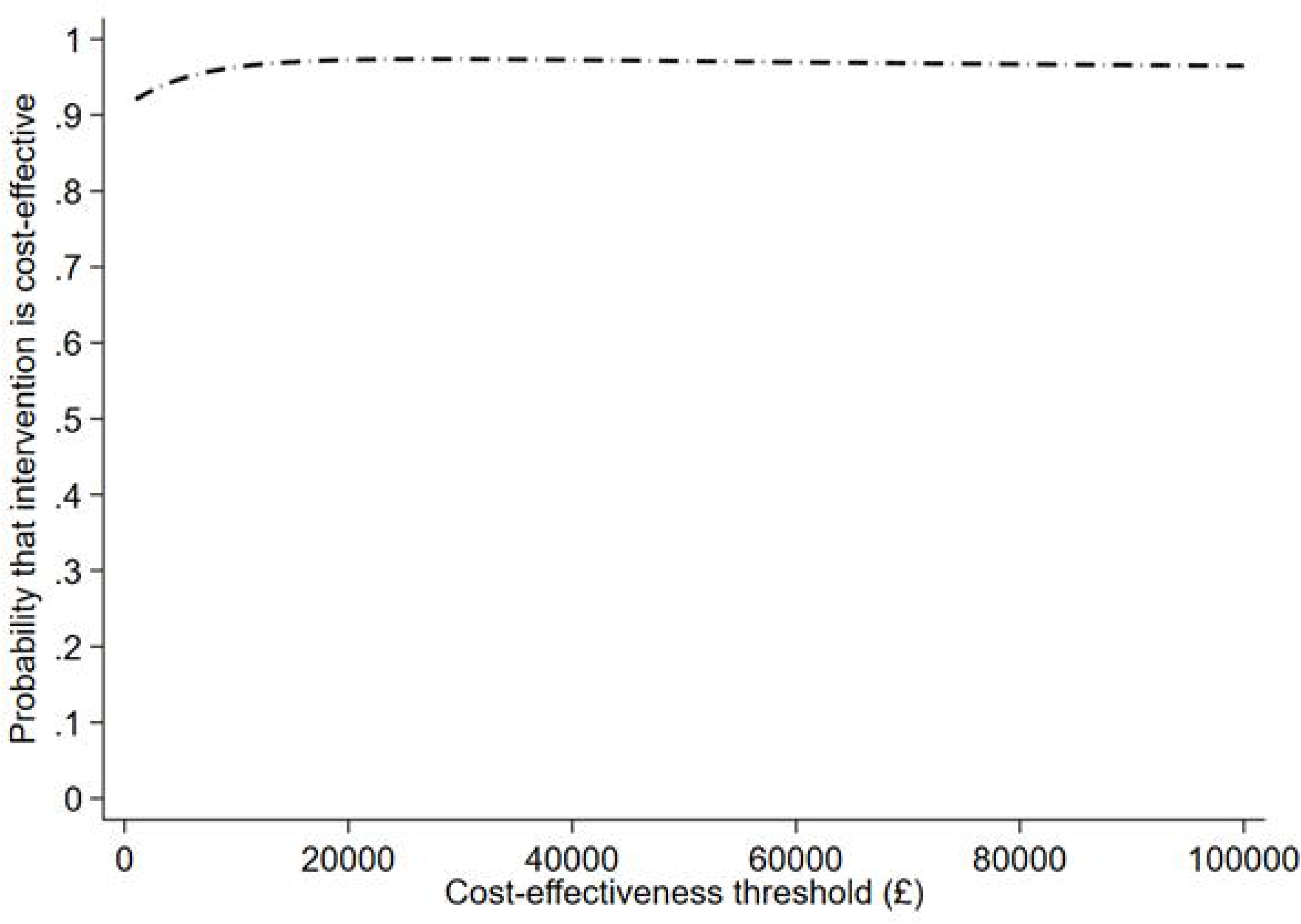

Cost effectiveness results were consistent from a societal perspective and in sensitivity analysis including only complete cases (Table 6). In a post-hoc sensitivity analysis including primary care costs only, results were marginally lower with a 95% probability of cost effectiveness at £20,000 per QALY. Adjusted mean years of full capability equivalent were higher in the intervention group, but confidence intervals spanned zero (0.003; 95% CI -0.011 to 0.017) (Supplementary Table 10). In the cost-effectiveness analysis, the intervention dominated usual care (i.e. lower costs and higher years of full capability equivalent). All outcomes and costs are tabulated in a cost-consequences table (Supplementary Table 11).

**Table 6.**
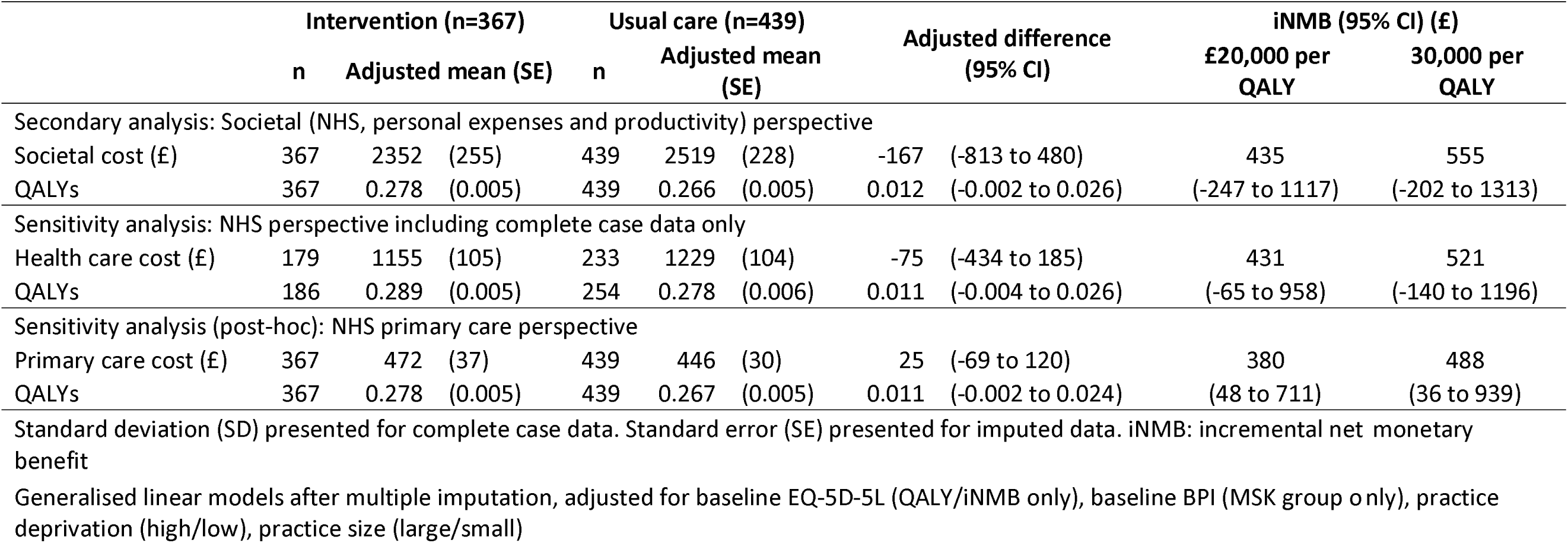
Secondary and sensitivity economic analyses, by intervention allocation for musculoskeletal pain group.

For the All-comers group, adjusted mean costs and QALYs were similar between arms (Supplementary Table 12). Mean resource use and costs are presented in Supplementary Table 13. From an NHS perspective, the INMB at a willingness-to-pay threshold of £20,000 was positive but small and not statistically significant (£134; 95% CI -£298 to £565), with a 73% probability of being cost effective. While adjusted mean costs were lower in the intervention group from an NHS perspective, from a societal perspective mean costs were higher, but confidence intervals indicated no evidence of a difference (£262; 95% CI -£296 to £820) (Supplementary Table 14). From a societal perspective, the INMB at £20,000 was lower at £87, with a lower probability that the intervention is cost effective (37%). Uncertainty is further presented in sensitivity analyses and a CEAC (Supplementary Figure 1). In the cost-effectiveness analysis, the intervention was less costly but resulted in fewer years of full capability equivalent; however, neither were statistically equivalent (Supplementary Table 10).

## DISCUSSION

This large, robust cluster-randomised trial found that a rigorously developed brief e-learning package on clinical empathy and realistic optimism for primary care practitioners produced a sustained increase in practitioner confidence to communicate clinical empathy and realistic optimism. We found no effect on patient-reported pain, enablement, satisfaction, perceptions of practitioner empathy and optimism, or other secondary outcomes. We also found no evidence of harm associated with the intervention. The novel economic evaluation of empathy training found that EMPathicO is a low-cost e-learning package to implement and has a high probability of being cost effective for patients with MSK pain. Cost effectiveness for other patients was less certain.

A key strength of this trial was the successful recruitment of a large sample of general practices (53), primary care practitioners from multiple professions (233), and patients (1682) from across England and Wales. Our sample was less ethnically diverse than the general population as indicated by the 2021 census: 5% of patient participants were from non-White ethnic minorities, compared to 18% of the general population and 10% of over 50s^77^. This was despite seeking practices that serve more diverse populations, recruiting 20% of practitioners from non-White ethnic minorities, preparing materials in multiple languages, offering interpreters, and working with a diverse group of PPI advisors. Blinding was maintained well, which can be challenging in trials of communication interventions: patients were not told (until after the trial) that the trial involved some practitioners undertaking training in communication skills, practitioners were not told which patients were participating in the trial, and in addition to the statisticians and lead researchers, all researchers involved in collecting patient reported outcomes were blinded to allocation. The economic evaluation provides new and comprehensive evidence for policy makers about the relative costs and benefits of EMPathicO. Cost-effectiveness results were consistent with primary analyses in a post-hoc sensitivity analysis exploring the impact of excluding secondary care costs, which was conducted due to high-cost outliers for secondary care, and a potential lag for secondary care referrals that result from primary care contacts during the follow-up period. However, the economic evaluation relied on self-report resource-use data, for which there was a large proportion of missing data. Evidence of differences in costs and outcomes was not observed.

In contrast to many other trials of empathy and similar communication training,^3 29^ we measured health outcomes (as well as patient satisfaction), practitioner self-efficacy, and patient-reported (rather than practitioner-reported) perceptions of clinical empathy. Other studies have similarly reported that practitioners find empathy training helpful but ours is the first to measure practitioner self-efficacy, an established determinant of behaviour change.^6^ ^29^ ^78^ The EMPathicO e-learning package incorporated content from a previous training called KEPe-WARM, which had significant effects on patient satisfaction.^32 79^ KEPe-WARM required practitioners to film and reflect on recordings of their consultations and was delivered face-to-face.^79^ In our feasibility study mandatory recording of consultations deterred trial participation and training uptake, hence we removed this requirement and only 3 practitioners chose to record their consultations in this trial.^42^ However, reflecting on recorded consultations may enhance behaviour change more than a brief, self-directed e-learning package. In-person delivery might be more effective than e-learning, but this is not observed in medical students learning communication skills^80^ and our practitioners found e-learning more feasible than in-person learning in the current pressurised primary care clinical context.^32^

The results indicate that the e-learning package confers sustained benefits to practitioners and is probably cost effective. Five possible explanations for the lack of effect on patient-reported outcomes warrant consideration. One, perhaps practitioners did not adopt EMPathicO techniques effectively. However, this is inconsistent with our data showing practitioners valued EMPathicO and described using their newly enhanced communication skills in consultations.^62^ Two, on average 38% of eligible practitioners in each practice took part, so effects may have been diluted as intervention arm patients consulted non-participating practitioners during follow-up. However, if that were the case we might expect differences between intervention and control at <7 day follow-up but not subsequently, whereas differences between intervention and control arms were broadly consistent over time. Three, there might have been ceiling effects on patient-perceived clinical empathy and realistic optimism, with high scores in both intervention and control arms. Average CARE^22^ scores ranged from 39.2 (intervention, MSK pain) to 42.5 (usual care, All-comers) out of a possible maximum of 50, which is broadly similar to the pooled mean CARE score of 40.5 (95% CI 39.2, 41.7) in a systematic review of 64 observational studies published up to 2016. While this comparison suggests ceiling effects were unlikely, they remain possible given the rapid changes in primary care since COVID-19 and the lack of more recent large-scale comparative data; practitioners who chose to take part in our trial may have already had better communication skills than those who chose not to. Four, the multifactorial nature of pain and enablement in primary care patients might have limited any effects of practitioner expressions of empathy and realistic optimism. Much (but not all) of the evidence for larger analgesic effects of positive messages derives from laboratory-based studies of induced pain or studies of acute pain.^24 81^ Primary care patients including those recruited into this large trial in NHS general practices may have symptoms that are less amenable to subtle changes in clinical empathy and realistic optimism within a single 10 minute consultation^82^ and often in the context of comorbidities.^83^ Five, our qualitative analysis suggests another, intriguing, explanation – that any immediate effects of patients’ positive experiences within consultations may have been diminished and rapidly truncated by the comparative lack of system level empathy experienced by patients when negotiating access to primary care appointments and subsequent referrals.^63 84^

Disseminating EMPathicO to primary care practitioners would be safe and economically viable and would enhance practitioner self-efficacy over at least 6 months. With widespread dissemination to trainees as well as qualified practitioners, EMPathicO could reach those with more scope to enhance their communication skills compared to the practitioners choosing to take part in this research trial. Given the current workforce crisis^85^ and the complexity of emerging links between empathy and practitioner burnout,^8^ future evaluations of empathy training should assess not only practitioner self-efficacy but also practitioner burnout and wellbeing. Further research could compare the acceptability and effects of the e-learning package with and without requiring practitioners to film their consultations. Research to develop and evaluate ways of enhancing the empathy of primary healthcare systems is also warranted.^86^ ^87^

## Supporting information

Consort cluster trials checklist

Consort abstracts checklist

TIDieR statement intervention description

## ADDITIONAL INFORMATION

### Registration

ISRCTN18010240 registered prospectively (before first patient enrolled) on 15 September 2022.

### Protocol

A paper presenting the trial protocol has been published;^37^ the full protocol and the statistical analysis plan are available in supplementary materials.

### Ethics Approval

Ethics approval was obtained from the South Central - Hampshire B Research Ethics Committee (ref: 22/SC/0145) and all participants gave informed consent before taking part.

### Transparency

The guarantor (F L Bishop) affirms that the manuscript is an honest, accurate, and transparent account of the study being reported; that no important aspects of the study have been omitted; and that any discrepancies from the study as originally planned and registered have been explained.

## Funding

Funding: This project was funded by the National Institute for Health Research (NIHR) School for Primary Care Research grant (project reference 563). The Primary Care Research Centre, University of Southampton is a member of the NIHR School for Primary Care Research and supported by NIHR Research funds. Service support costs will be paid by the CRN. CDM is funded by the National Institute for Health Research (NIHR) Collaborations for Leadership in Applied Health Research and Care West Midlands and the NIHR School for Primary Care Research. The EMPathicO e-learning package was developed using LifeGuide software, which was partly funded by the National Institute for Health Research Southampton Biomedical Research Centre (BRC). NIHR Local Clinical Research Networks (CRNs) supported practice recruitment.

The views expressed are those of the authors and not necessarily those of the NIHR or the Department of Health and Social Care.

The study sponsor (University of Southampton) and funders had no role in study design; collection, management, analysis, and interpretation of data; writing of the report; or the decision to submit the report for publication. The researchers are independent from the funders and all authors had full access to all of the data (including statistical reports and tables) in the study and can take responsibility for the integrity of the data and the accuracy of the data analysis.

## Acknowledgements

The TIP trial team gratefully acknowledge the contribution of our Public Advisory Group (Jennifer Bostock, Clara Martins de Barros, Mark Lamond, Manoj Mistry, Hazel Patel, and David Truswell), our Independent Trial Steering Committee (Joanne Reeve, Ian Dickerson, Ines Rombach, Philip Pallman), and our administrator (Tanya Palmer).

## Contributor and Guarantor Information

All authors meet the ICMJE criteria for authorship, i.e., have made substantial contributions to the conception or design of the work; or the acquisition, analysis, or interpretation of data for the work; AND drafted the work or reviewed it critically for important intellectual content; AND approved the version to be published; AND agree to be accountable for all aspects of the work in ensuring that questions related to the accuracy or integrity of any part of the work are appropriately investigated and resolved.

Specific contributions as per the CRediT taxonomy are: Conceptualization (FLB, TB, KG, NC, RDH, ET, AH, MER, MJR, CM, LC, JB, BS, LM, SP, JV, HA, JH, GML, JN, NI, PHL, PL, HAE); Data curation (TB, KG, NC); Formal analysis (TB, KG); Funding acquisition (FLB, KG, MJR, CM, LC, JB, BS, LM, JV, HA, JH, GML, PL, HAE), Investigation (NC, RDH, ET, AH, MER, SP, NI), Methodology (FLB, TB, KG, PL, HAE), Project administration (FLB, NC, HAE, JN), Software (SP), Supervision (FLB, MJR, CM, LC, JN, HAE), Validation (BS), Writing – original draft (FLB, TB, KG), Writing – review & editing (FLB, TB, KG, NC, RDH, ET, AH, MER, MJR, CM, LC, JB, BS, LM, SP, JV, HA, JH, GML, JN, NI, PHL, PL, HAE).

The guarantor (F L Bishop) accepts full responsibility for the work and/or the conduct of the study, had access to the data, and controlled the decision to publish. The corresponding author attests that all listed authors meet authorship criteria and that no others meeting the criteria have been omitted.

## Copyright/License for Publication

The Corresponding Author has the right to grant on behalf of all authors and does grant on behalf of all authors, a worldwide licence to the Publishers and its licensees in perpetuity, in all forms, formats and media (whether known now or created in the future), to i) publish, reproduce, distribute, display and store the Contribution, ii) translate the Contribution into other languages, create adaptations, reprints, include within collections and create summaries, extracts and/or, abstracts of the Contribution, iii) create any other derivative work(s) based on the Contribution, iv) to exploit all subsidiary rights in the Contribution, v) the inclusion of electronic links from the Contribution to third party material where-ever it may be located; and, vi) licence any third party to do any or all of the above.

## Declaration of Competing Interest

The authors declare the following competing interests: FLB (research grant from NIHR School for Primary Care Research paid to institution; speakers honoraria from Stoneygate Centre for Empathic Healthcare and New Scientist), TB (research grant from NIHR School for Primary Care Research paid to institution), KG (research grant from NIHR School for Primary Care Research paid to institution; ModRUM license holder), NC (research grant from NIHR School for Primary Care Research paid to institution), RDH (none declared), ET (research grant from NIHR School for Primary Care Research paid to institution), AH (research grant from NIHR School for Primary Care Research paid to institution), MER (research grant from NIHR School for Primary Care Research paid to institution), MJR (research grant from NIHR School for Primary Care Research paid to institution; other research funding from NIHR SPCR, HTA, and PGfAR paid to institution; NIHR Research Professorship; on TSC/DMC for ERICA, BabyBathe and ASYMPTOMATIC trials), CM (research grant from NIHR School for Primary Care Research paid to institution; other funding from NIHR, MRC, NHS paid to institution; is the Director of the NIHR SPCR), LC (research grant from NIHR School for Primary Care Research paid to institution; paid role as Chief Medical Officer, NHS Shropshire, Telford and Wrekin; Salaried GP at Brook Medical Centre, Stoke-on-Trent; Senior Lecturer in General Practice Research, Keele University), JB (research grant from NIHR School for Primary Care Research), BS (research grant from NIHR School for Primary Care Research paid to institution; other research funding from NIHR paid to institution; member of NIHR HTA Commissioning Panel – 15/09/2020 to present), LM (research grant from NIHR School for Primary Care Research paid to institution), SP (research grant from NIHR School for Primary Care Research paid to institution), JV (none declared), HA (research grant from NIHR School for Primary Care Research paid to institution; other research funding from NIHR, Research Council of Norway, University of Warwick/eConsult Ltd paid to institution; honoraria from Imperial College London, UCL, North West Cancer Research; received support for travel from NIHR, RCGP, University of Birmingham; advisory board member/chair for NIHR159467, NIHR160384, BRACE rapid evaluation centre, HED-LINE study, EPaCCS study; is Vice Chair of the Scientific Foundation Board, Royal College of General Practitioners), JH (research grant from NIHR School for Primary Care Research paid to institution), GML (none declared), JN (none declared), NI (none declared), PHL (none declared), PL (research grant from NIHR School for Primary Care Research paid to institution), HAE (research grant from NIHR School for Primary Care Research paid to institution; is Deputy Academic Capacity Development Lead for the NIHR SPCR and sits on the NIHR SPCR Board and Exec; works clinically as a GP at New Horizons Medical Partnership in Southampton as part of Professor of Primary Care Research post at the University of Southampton; Co-Authors the Oxford Handbook of General Practice published by Oxford University Press).

## Notes

### Clinical Trial

ISRCTN18010240

### Clinical Protocols

https://bmjopen.bmj.com/content/14/3/e081932

### Author Declarations

Ethic committee South Central - Hampshire B Research Ethics Committee gave ethical approval for this work (reference: 22/SC/0145).

## References

1. Howick J, Bizzari V, Dambha-Miller H. Therapeutic empathy: what it is and what it isn’t. J R Soc Med 2018;111(7):233–36. doi: 10.1177/0141076818781403

2. Derksen F, Bensing J, Lagro-Janssen A. Effectiveness of empathy in general practice: a systematic review. Br J Gen Pract2013;63(606):e76-84. doi: 10.3399/bjgp13X660814 [published Online First: 2013/01/23]

3. Byrne M, Campos C, Daly S, et al. The current state of empathy, compassion and person-centred communication training in healthcare: An umbrella review. Patient Educ Couns 2024;119:108063. doi: 10.1016/j.pec.2023.108063 [published Online First: 2023/11/27]

4. Keshtkar L, Madigan CD, Ward A, et al. The Effect of Practitioner Empathy on Patient Satisfaction : A Systematic Review of Randomized Trials. Ann Intern Med 2024;177(2):196–209. doi: 10.7326/m23-2168 [published Online First: 2024/01/29]

5. Howick J, Moscrop A, Mebius A, et al. Effects of empathic and positive communication in healthcare consultations: a systematic review and meta-analysis. J R Soc Med 2018;111(7):240–52. doi: 10.1177/0141076818769477 [published Online First: 2018/04/20]

6. Winter R, Leanage N, Roberts N, et al. Experiences of empathy training in healthcare: A systematic review of qualitative studies. Patient Educ Couns 2022;105(10):3017–37. doi: 10.1016/j.pec.2022.06.015 [published Online First: 2022/07/11]

7. Wilkinson H, Whittington R, Perry L, et al. Examining the relationship between burnout and empathy in healthcare professionals: A systematic review. Burn Res 2017;6:18–29. doi: 10.1016/j.burn.2017.06.003 [published Online First: 2017/09/05]

8. Delgado N, Delgado J, Betancort M, et al. What is the Link Between Different Components of Empathy and Burnout in Healthcare Professionals? A Systematic Review and Meta-Analysis. Psychol Res Behav Manag 2023;16:447–63. doi: 10.2147/prbm.S384247 [published Online First: 2023/02/24]

9. Cairns P, Isham AE, Zachariae R. The association between empathy and burnout in medical students: a systematic review and meta-analysis. BMC Med Educ 2024;24(1):640. doi: 10.1186/s12909-024-05625-6 [published Online First: 2024/06/08]

10. Thorne SE, Bultz BD, Baile WF. Is there a cost to poor communication in cancer care?: a critical review of the literature. Psychooncology 2005;14(10):875–84. doi: 10.1002/pon.947

11. Barry CA, Bradley CP, Britten N, et al. Patients’ unvoiced agendas in general practice consultations: qualitative study. Br Med J 2000;320:1246–50.

12. Pincock S. Poor communication lies at heart of NHS complaints, says ombudsman. BMJ 2004;328(7430):10.

13. Stelfox HT, Gandhi TK, Orav EJ, et al. The relation of patient satisfaction with complaints against physicians and malpractice lawsuits. The American Journal of Medicine 2005;118(10):1126–33. doi: 10.1016/j.amjmed.2005.01.060

14. Gilligan C, Powell M, Lynagh MC, et al. Interventions for improving medical students’ interpersonal communication in medical consultations. Cochrane Database of Systematic Reviews 2021(2) doi: 10.1002/14651858.CD012418.pub2

15. Engbers RA. Students’ perceptions of interventions designed to foster empathy: An integrative review. Nurse Educ Today 2020;86:104325. doi: 10.1016/j.nedt.2019.104325 [published Online First: 2020/01/12]

16. Menezes P, Guraya SY, Guraya SS. A Systematic Review of Educational Interventions and Their Impact on Empathy and Compassion of Undergraduate Medical Students. Front Med (Lausanne) 2021;8:758377. doi: 10.3389/fmed.2021.758377 [published Online First: 2021/11/26]

17. Gong B, Zhang X, Lu C, et al. The effectiveness of Balint groups at improving empathy in medical and nursing education: a systematic review and meta-analysis of randomized controlled trials. BMC Med Educ 2024;24(1):1089. doi: 10.1186/s12909-024-06098-3

18. Cho MK, Kim MY. Effectiveness of simulation-based interventions on empathy enhancement among nursing students: a systematic literature review and meta-analysis. BMC Nurs 2024;23(1):319. doi: 10.1186/s12912-024-01944-7 [published Online First: 2024/05/12]

19. Zhang X, Pang HF, Duan Z. Educational efficacy of medical humanities in empathy of medical students and healthcare professionals: a systematic review and meta-analysis. BMC Med Educ 2023;23(1):925. doi: 10.1186/s12909-023-04932-8 [published Online First: 2023/12/07]

20. Vennik J, Hughes S, Smith KA, et al. Patient and practitioner priorities and concerns about primary healthcare interactions for osteoarthritis: A meta-ethnography. Patient Educ Couns 2022 doi: 10.1016/j.pec.2022.01.009

21. Howick J, Steinkopf L, Ulyte A, et al. How empathic is your healthcare practitioner? A systematic review and meta-analysis of patient surveys. BMC Med Educ 2017;17(1):136. doi: 10.1186/s12909-017-0967-3 [published Online First: 2017/08/22]

22. Mercer SW, Maxwell M, Heaney D, et al. The development and preliminary validation of the Consultation and Relational Empathy (CARE) measure: an empathy-based consultation process measure. Fam Pract 2004;21 699–705.

23. Hughes S, Vennik JL, Smith KA, et al. Clinician views on optimism and empathy in primary care consultations: a qualitative interview study. BJGP Open 2022;6(3):BJGPO.2021.0221. doi: 10.3399/BJGPO.2021.0221

24. Colloca L. The Placebo Effect in Pain Therapies. Annu Rev Pharmacol Toxicol 2019;59:191–211. doi: 10.1146/annurev-pharmtox-010818-021542 [published Online First: 2018/09/14]

25. Benedetti F, Frisaldi E, Shaibani A. Thirty Years of Neuroscientific Investigation of Placebo and Nocebo: The Interesting, the Good, and the Bad. Annu Rev Pharmacol Toxico2l 022;62:323–40. doi: 10.1146/annurev-pharmtox-052120-104536 [published Online First: 2021/08/31]

26. Kaptchuk TJ, Shaw J, Kerr CE, et al. “Maybe I made up the whole thing”: Placebos and patients’ experiences in a randomized controlled trial. Cult Med Psychiatry 2009;33:382–411.

27. Hyland ME. Motivation and placebos: do different mechanisms occur in different contexts? Philosophical Transactions of the Royal Society B: Biological Sciences 2011;366(1572):1828–37.

28. Evers AWM, Colloca L, Blease C, et al. Implications of Placebo and Nocebo Effects for Clinical Practice: Expert Consensus. Psychother Psychosom 2018;87(4):204–10. doi: 10.1159/000490354 [published Online First: 2018/06/12]

29. Winter R, Issa E, Roberts N, et al. Assessing the effect of empathy-enhancing interventions in health education and training: a systematic review of randomised controlled trials. BMJ Open 2020;10(9):e036471. doi: 10.1136/bmjopen-2019-036471 [published Online First: 2020/09/27]

30. Smith KA, Bishop FL, Dambha-Miller H, et al. Improving Empathy in Healthcare Consultations-a Secondary Analysis of Interventions. J Gen Intern Med 2020;35(10):3007–14. doi: 10.1007/s11606-020-05994-w [published Online First: 2020/07/14]

31. Howick J, Mittoo S, Abel L, et al. A price tag on clinical empathy? Factors influencing its cost-effectiveness. J R Soc Med 2020;113(10):389–93. doi: 10.1177/0141076820945272

32. Smith KA, Vennik J, Morrison L, et al. Harnessing placebo effects in primary care: Using the person-based approach to develop an online intervention to enhance practitioners’ communication of clinical empathy and realistic optimism during consultations. Frontiers in Pain Research2021;2 doi: 10.3389/fpain.2021.721222

33. Howick J, Lyness E, Albury C, et al. Anatomy of positive messages in healthcare consultations: component analysis of messages within 22 randomised trials. Eur J Pers Cent Healthc 2019;17:656–64.

34. Budd G, Griffiths D, Howick J, et al. Empathy in patient-clinician interactions when using telecommunication: A rapid review of the evidence. PEC Innovation 2022;1:100065. doi: 10.1016/j.pecinn.2022.100065

35. Lyness E, Vennik JL, Bishop FL, et al. Exploring patient views of empathic optimistic communication for osteoarthritis in primary care: a qualitative interview study using vignettes. BJGP Open 2021:BJGPO.2021.0014. doi: 10.3399/BJGPO.2021.0014

36. Vennik J, Hughes S, Lyness E, et al. Patient perceptions of empathy in primary care telephone consultations: A mixed methods study. Patient Educ Couns 2023;113:107748. doi: 10.1016/j.pec.2023.107748

37. Bishop FL, Cross N, Dewar-Haggart R, et al. Talking in primary care (TIP): protocol for a cluster-randomised controlled trial in UK primary care to assess clinical and cost-effectiveness of communication skills e-learning for practitioners on patients’ musculoskeletal pain and enablement. BMJ Open 2024;14(3):e081932. doi: 10.1136/bmjopen-2023-081932

38. Gill TK, Mittinty MM, March LM, et al. Global, regional, and national burden of other musculoskeletal disorders, 1990&#x2013;2020, and projections to 2050: a systematic analysis of the Global Burden of Disease Study 2021. The Lancet Rheumatology 2023;5(11):e670-e82. doi: 10.1016/S2665-9913(23)00232-1

39. Organization WH. International Statistical Classification of Diseases and Related Health Problems (ICD). https://www.who.int/standards/classifications/classification-of-diseases, 2021.

40. Keller S, Bann CM, Dodd SL, et al. Validity of the Brief Pain Inventory for use in documenting the outcomes of patients with noncancer pain. Clin J Pain 2004;20(5):309–18.

41. Whitaker P. Ticking the ICE box: the future of doctor-patient communication in a post-covid world. BMJ 2021;373:n870. doi: 10.1136/bmj.n870

42. Bishop FL, Howick J, Vennik J, et al. Feasibility trial of a new digital training package to enhance primary care practitioners’ communication of clinical empathy and realistic optimism, 2025 [Under Review].

43. Kreuter MW, Wray RJ. Tailored and targeted health communication: strategies for enhancing information relevance. American Journal of Health Behavior2003;27(1):S227–S32.

44. LifeGuide+ 2025 [Available from: https://lifeguideonline.org/ accessed 3rd February 2025.

45. Howie JG, Heaney DJ, Maxwell M, et al. A comparison of a Patient Enablement Instrument (PEI) against two established satisfaction scales as an outcome measure of primary care consultations. Fam Pract 1998;15(2):165–71.

46. Morrison LG, Geraghty AWA, Lloyd S, et al. Comparing usage of a web and app stress management intervention: An observational study. Internet Interventions 2018;12:74–82. doi: 10.1016/j.invent.2018.03.006

47. Molgaard Nielsen A, Hartvigsen J, Kongsted A, et al. The patient enablement instrument for back pain: reliability, content validity, construct validity and responsiveness. Health and quality of life outcomes2021;19(1):116. doi: 10.1186/s12955-021-01758-0 [published Online First: 2021/04/11]

48. Bedford LE, Yeung MHY, Au CH, et al. The validity, reliability, sensitivity and responsiveness of a modified Patient Enablement Instrument (PEI-2) as a tool for serial measurements of health enablement. Fam Pract 2020 doi: 10.1093/fampra/cmaa102

49. Preston CC, Colman AM. Optimal number of response categories in rating scales: reliability, validity, discriminating power, and respondent preferences. Acta Psychol (Amst) 2000;104(1):1–15. doi: 10.1016/S0001-6918(99)00050-5

50. Fischer D, Stewart AL, Bloch DA, et al. Capturing the Patient’s View of Change as a Clinical Outcome Measure. JAMA 1999;282(12):1157–62. doi: 10.1001/jama.282.12.1157

51. Wolf MH, Putnam SM, James SA, et al. The Medical Interview Satisfaction Scale: development of a scale to measure patient perceptions of physician behavior. J Behav Med 1978;1(4):391–401. [published Online First: 1978/12/01]

52. Meakin R, Weinman J. The ‘Medical Interview Satisfaction Scale’ (MISS-21) adapted for British general practice. Fam Pract 2002;19(3):257–63. doi: 10.1093/fampra/19.3.257

53. Herdman M, Gudex C, Lloyd A, et al. Development and preliminary testing of the new five-level version of EQ-5D (EQ-5D-5L). Qual Life Res 2011;20(10):1727–36. doi: 10.1007/s11136-011-9903-x [published Online First: 2011/04/12]

54. Al-Janabi H, Flynn TN, Coast J. Development of a self-report measure of capability wellbeing for adults: the ICECAP-A. Qual Life Res 2012;21(1):167–76. doi: 10.1007/s11136-011-9927-2 [published Online First: 2011/05/21]

55. Keeley T, Coast J, Nicholls E, et al. An analysis of the complementarity of ICECAP-A and EQ-5D-3 L in an adult population of patients with knee pain. Health and quality of life outcomes 2016;14:36. doi: 10.1186/s12955-016-0430-x [published Online First: 2016/03/05]

56. Garfield K, Husbands S, Thorn JC, et al. Development of a brief, generic, modular resource-use measure (ModRUM): cognitive interviews with patients. BMC Health Serv Res 2021;21(1):371. doi: 10.1186/s12913-021-06364-w

57. Reilly MC, Zbrozek AS, Dukes EM. The validity and reproducibility of a work productivity and activity impairment instrument. Pharmacoeconomics 1993;4(5):353–65. doi: 10.2165/00019053-199304050-00006 [published Online First: 1993/10/05]

58. Alberts J, Löwe B, Glahn MA, et al. Development of the generic, multidimensional Treatment Expectation Questionnaire (TEX-Q) through systematic literature review, expert surveys and qualitative interviews. BMJ Open 2020;10(8):e036169. doi: 10.1136/bmjopen-2019-036169

59. Bjelland I, Dahl AA, Haug TT, et al. The validity of the Hospital Anxiety and Depression Scale: An updated literature review. J Psychosom Res 2002;52(2):69–77.

60. Zigmond AS, Snaith RP. The Hospital Anxiety and Depression Scale. Acta Psychiatr Scand 1983;67(6):361–70.

61. Ridd MJ, Lewis G, Peters TJ, et al. Patient-Doctor Depth-of-Relationship Scale: Development and Validation. The Annals of Family Medicine 2011;9(6):538. doi: 10.1370/afm.1322

62. Teasdale E, Dewar-Haggart R, Pollet S, et al. Primary care practitioners’ views and experiences of undertaking and implementing ‘EMPathicO’ communication skills e-learning in everyday clinical practice: a nested qualitative study., 2025 [in submission].

63. Dewar-Haggart R, Teasdale E, Pollet S, et al. Patients’ perceptions of primary care consultations: is empathy under threat? A nested qualitative study [paper presentation]. Society for Academic Primary Care (SAPC) South West Conference. Oxford, UK, 2025.

64. Dworkin RH, Turk DC, Wyrwich KW, et al. Interpreting the clinical importance of treatment outcomes in chronic pain clinical trials: IMMPACT recommendations. J Pain 2008;9(2):105–21. doi: 10.1016/j.jpain.2007.09.005 [published Online First: 2007/12/07]

65. Adams G, Gulliford MC, Ukoumunne OC, et al. Patterns of intra-cluster correlation from primary care research to inform study design and analysis. J Clin Epidemiol 2004;57(8):785–94. doi: 10.1016/j.jclinepi.2003.12.013 [published Online First: 2004/10/16]

66. Bishop FL, Smith KA, Vennik J, et al. Feasibility Study of a Novel Online Intervention to Enhance Practitioners’ Communication of Clinical Empathy and Realistic Optimism During Primary Care Consultations, 2024, in preparation.

67. Hernandez Alava M, Pudney S, Wailoo A. Estimating the relationship between EQ-5D-5L and EQ-5D-3L: Results from an English population study. : Universities of Sheffield and York. Contract No. 063., 2000.

68. Hernandez Alava M, Pudney S. eq5dmap: A command for mapping between EQ-5D-3L and EQ-5D-5L. The Stata Journal 2018;18(2):395–415.

69. National Institute for Health and Care Excellence. NICE health technology evaluations: the manual 2022 doi: https://www.nice.org.uk/process/pmg36/resources/nice-health-technology-evaluations-the-manual-pdf-72286779244741

70. Manca A, Hawkins N, Sculpher MJ. Estimating mean QALYs in trial-based cost-effectiveness analysis: the importance of controlling for baseline utility. Health Econ 2005;14(5):487–96. doi: 10.1002/hec.944 [published Online First: 2004/10/22]

71. Jones K, Weatherly H, Birch S, et al. Unit Costs of Health and Social Care 2023 Manual. Technical report. Kent, UK, 2024.

72. NHS England. National Schedule of NHS Costs 2022/23, 2024.

73. NHS Business Services Authority. Prescription Cost Analysis - England - 2022/23, 2023.

74. Office for National Statistics. Annual Survey of Hours and Earnings (ASHE) 2023.

75. NHS England and NHS Improvement. National Schedule of NHS Costs, 2020.

76. Joint Formulary Committee. British National Formulary (online). London: BMJ Group and Pharmaceutical Press.

77. Office for National Statistics (ONS). Ethnic group by age and sex, England and Wales: Census 2021. https://www.ons.gov.uk/peoplepopulationandcommunity/culturalidentity/ethnicity/articles/ethnicgroupbyageandsexenglandandwales/census2021: ONS website, released 23 January 2023.

78. Sheeran P, Maki A, Montanaro E, et al. The impact of changing attitudes, norms, and self-efficacy on health-related intentions and behavior: A meta-analysis. Health Psychol 2016;35(11):1178–88. doi: 10.1037/hea0000387 [published Online First: 2016/10/18]

79. Little P, White P, Kelly J, et al. Randomised controlled trial of a brief intervention targeting predominantly non-verbal communication in general practice consultations. Br J Gen Pract 2015;65(635):e351–e56. doi: 10.3399/bjgp15X685237

80. Kyaw BM, Posadzki P, Paddock S, et al. Effectiveness of Digital Education on Communication Skills Among Medical Students: Systematic Review and Meta-Analysis by the Digital Health Education Collaboration. J Med Internet Res 2019;21(8):e12967. doi: 10.2196/12967

81. Mistiaen P, van Osch M, van Vliet L, et al. The effect of patient-practitioner communication on pain: a systematic review. European Journal of Pain 2016;20(5):675–88. doi: 10.1002/ejp.797 [published Online First: 2015/10/23]

82. Irving G, Neves AL, Dambha-Miller H, et al. International variations in primary care physician consultation time: a systematic review of 67 countries. BMJ Open 2017;7(10):e017902. doi: 10.1136/bmjopen-2017-017902

83. MacRae C, Mercer SW, Henderson D, et al. Age, sex, and socioeconomic differences in multimorbidity measured in four ways: UK primary care cross-sectional analysis. Br J Gen Pract 2023;73(729):e249. doi: 10.3399/BJGP.2022.0405

84. Wise J. Patient satisfaction in GP services falls sharply in latest survey. BMJ 2022;378:o1764. doi: 10.1136/bmj.o1764

85. Jefferson L, Holmes M. GP workforce crisis: what can we do now? Br J Gen Pract 2022;72(718):206. doi: 10.3399/bjgp22X719225

86. Kerasidou A, Bærøe K, Berger Z, et al. The need for empathetic healthcare systems. J Med Ethics 2021;47(12):e27. doi: 10.1136/medethics-2019-105921

87. Howick J, de Zulueta P, Gray M. Beyond empathy training for practitioners: Cultivating empathic healthcare systems and leadership. J Eval Clin Pract 2024;30(4):548–58. doi: 10.1111/jep.13970

