## Supplementary Tables for "Clinical and cost-effectiveness of communication skills e-learning for primary care practitioners on patients’ musculoskeletal pain and enablement: the Talking in Primary care (TIP) cluster-randomised controlled trial"

Supplementary Tables and Figures

### Supplementary Table 1. Baseline Characteristics of Patients in the All-Comers Group

|  | Usual care | Intervention | Total |
| --- | --- | --- | --- |
|  | n(%) | n(%) | n(%) |
| Participants, n | 386 | 490 | 876 |
| Age |  |  |  |
| median (IQR) | 60 (47-69) | 59 (45-70) | 60 (46-70) |
| mean (SD) | 56.9 (15.6) | 57.0 (15.9) | 57.0 (15.8) |
| range | 18-91 | 18-89 | 18-91 |
| missing | 1 | 1 | 2 |
| Gender |  |  |  |
| Male | 145 (37.6) | 178 (36.3) | 323 (36.9) |
| Female | 240 (62.2) | 311 (63.5) | 551 (62.9) |
| Prefer not to say | 0 (0.0) | 1 (0.2) | 1 (0.1) |
| Other | 1 (0.3) | 0 (0.0) | 1 (0.1) |
| Ethnicity |  |  |  |
| White | 368 (95.3) | 463 (94.5) | 831 (94.9) |
| Mixed / Multiple ethnic groups | 7 (1.8) | 8 (1.6) | 15 (1.7) |
| Asian / Asian British | 4 (1.0) | 9 (1.8) | 13 (1.5) |
| Black/ African / Caribbean / Black British | 3 (0.8) | 6 (1.2) | 9 (1.0) |
| Other | 4 (1.0) | 4 (0.8) | 8 (0.9) |
| Educational level |  |  |  |
| No formal educational qualifications | 24 (6.2) | 37 (7.6) | 61 (7.0) |
| GCSEs / O levels or similar | 86 (22.3) | 100 (20.5) | 186 (21.3) |
| A levels or similar or ONC / OND | 37 (9.6) | 75 (15.4) | 112 (12.8) |
| HNC / HND Degree | 40 (10.4) | 33 (6.8) | 73 (8.4) |
| Degree | 107 (27.7) | 124 (25.4) | 231 (26.4) |
| Higher degree | 18 (4.7) | 29 (5.9) | 47 (5.4) |
| Postgraduate degree | 53 (13.7) | 69 (14.1) | 122 (14.0) |
| Other | 21 (5.4) | 21 (4.3) | 42 (4.8) |
| Missing | 0 (0) | 2 (0.0) | 2 (0.0) |
| Employment |  |  |  |
| Employed full-time | 128 (33.2) | 144 (29.4) | 272 (31.1) |
| Employed part-time | 48 (12.5) | 64 (13.1) | 112 (12.8) |
| Retired | 143 (37.1) | 199 (40.6) | 342 (39.1) |
| Unemployed | 7 (1.8) | 12 (2.4) | 19 (2.2) |
| Doing unpaid work (e.g. volunteering) | 6 (1.6) | 5 (1.0) | 11 (1.3) |
| Unable to work | 24 (6.2) | 32 (6.5) | 56 (6.4) |
| Other | 29 (7.5) | 34 (6.9) | 63 (7.2) |
| Missing | 1 (0.0) | 0 (0) | 1 (0.0) |
| Index multiple deprivation, median (IQR) | 7 (5-9) | 6 (4-8) | 7 (4-9) |
| Pain intensity last week (BPI) n | 176 | 214 | 390 |
| Pain intensity last week (BPI), mean (SD) | 3.7 (2.2) | 3.8 (2.2) | 3.7 (2.2) |
| Pain intensity subscale (BPI), n | 177 | 216 | 393 |
| Pain intensity subscale (BPI), mean (SD) | 3.6 (2.1) | 3.6 (2.1) | 3.6 (2.1) |
| Symptom severity (PGIS), mean (SD) | 3.2 (1.4) | 3.3 (1.5) | 3.2 (1.5) |
| Symptom severity (PGIS), missing | 1 (0.0) | 0 (0) | 1 (0.0) |
| Main reason for consulting |  |  |  |
| General | 85 (22.0) | 111 (22.7) | 195 (22.3) |
| Blood, blood-forming organs, immune system | 4 (1.0) | 4 (0.8) | 8 (0.9) |
| Digestive system | 20 (5.2) | 33 (6.7) | 53 (6.1) |
| Eye | 2 (0.5) | 2 (0.4) | 4 (0.5) |
| Genital system | 18 (4.7) | 25 (5.1) | 43 (4.9) |
| Ear | 5 (1.3) | 9 (1.8) | 14 (1.6) |
| Interventions and processes | 29 (7.5) | 22 (4.5) | 51 (5.8) |
| Functioning and functioning-related | 3 (0.8) | 4 (0.8) | 7 (0.8) |
| Circulatory system | 20 (5.2) | 36 (7.4) | 56 (6.4) |
| Musculoskeletal | 67 (17.4) | 58 (11.8) | 125 (14.3) |
| Neurological system | 16 (4.2) | 22 (4.5) | 38 (4.3) |
| Psychological, mental, neurodevelopmental | 20 (5.2) | 30 (6.1) | 50 (5.7) |
| Respiratory | 32 (8.3) | 46 (9.4) | 78 (8.9) |
| Skin | 29 (7.5) | 37 (7.6) | 66 (7.5) |
| Endocrine, metabolic, nutritional system | 20 (5.2) | 31 (6.3) | 51 (5.8) |
| Urinary system | 9 (2.3) | 13 (2.7) | 22 (2.5) |
| Pregnancy and childbearing | 7 (1.8) | 7 (1.4) | 14 (1.6) |
| Appointment modality |  |  |  |
| In person | 285 (73.8) | 323 (65.9) | 608 (69.4) |
| Telephone | 101 (26.2) | 165 (33.7) | 266 (30.4) |
| Video | 0 (0.0) | 2 (0.4) | 2 (0.2) |

BPI Brief Pain Inventory on a scale 0 (no pain) to 10 (most pain); Patient Global Impression of Severity on scale 1 (none) to 7 (extremely severe). Main reason for consulting coded according to ICPC-3 Chapter.

### Supplementary Table 2. Self-Reported Pain Medication Use in the MSK Pain Group, by Intervention Allocation

| Pain medication | Usual care  (N=439) | Intervention  (N=367) |
| --- | --- | --- |
| 7 days, n | 327 | 260 |
| Paracetamol | 121 (37.0%) | 93 (35.8%) |
| Ibuprofen | 75 (22.9%) | 61 (23.5%) |
| Ibuprofen gel | 11 (3.4%) | 3 (1.2%) |
| 3 months, n | 291 | 212 |
| Paracetamol | 123 (42.3%) | 92 (43.4%) |
| Ibuprofen | 63 (21.7%) | 52 (24.5%) |
| Ibuprofen gel | 10 (3.4%) | 5 (2.4%) |
| 6 months, n | 275 | 214 |
| Paracetamol | 107 (38.9%) | 89 (41.6%) |
| Ibuprofen | 45 (16.4%) | 48 (22.3%) |
| Ibuprofen gel | 9 (3.3%) | 9 (4.2%) |

### Supplementary Table 3. All-Comers Group Primary and Secondary Outcomes by Intervention Allocation

|  | Usual care (N=386) | | Intervention (N=490) | | Mean difference |
| --- | --- | --- | --- | --- | --- |
|  | n | Mean (SD) | n | Mean (SD) | (95% CI) |
| **Primary Outcomes** |  |  |  |  |  |
| **Patient enablement (PEI)** |  |  |  |  |  |
| 7 days | 234 | 5.1 (1.2) | 299 | 5.0 (1.3) |  |
| 1 month | 233 | 5.0 (1.1) | 297 | 4.7 (1.3) |  |
| 3 months | 221 | 5.0 (1.0) | 280 | 5.0 (1.2) |  |
| 6 months | 205 | 5.1 (1.1) | 279 | 5.0 (1.3) |  |
| Repeated measures over 6 months | 316 |  | 397 |  | -0.12 (-0.32, 0.07) |
| **Secondary Outcomes** |  |  |  |  |  |
| **Symptom severity (PGIS)** |  |  |  |  |  |
| 7 days | 282 | 3.0 (1.3) | 367 | 3.0 (1.4) |  |
| 1 month | 264 | 2.8 (1.4) | 341 | 3.0 (1.5) |  |
| 3 months | 247 | 2.7 (1.4) | 310 | 2.8 (1.5) |  |
| 6 months | 229 | 2.9 (1.5) | 299 | 2.9 (1.6) |  |
| Repeated measures | 330 |  | 411 |  | 0.05 (-0.13, 0.24) |
| **Symptom change (PGIC)** |  |  |  |  |  |
| 7 days | 272 | 3.5 (0.9) | 356 | 3.6 (0.9) |  |
| 1 month | 253 | 3.2 (1.2) | 322 | 3.4 (1.2) |  |
| 3 months | 246 | 3.1 (1.2) | 309 | 3.2 (1.3) |  |
| 6 months | 224 | 3.2 (1.3) | 285 | 3.2 (1.3) |  |
| Repeated measures | 327 |  | 408 |  | 0.08 (-0.04, 0.21) |
| **Patient satisfaction (MISS UK)** | 272 | 5.3 (0.8) | 353 | 5.3 (0.9) | -0.09 (-0.26, 0.08) |
| Distress relief | 274 | 4.6 (1.1) | 359 | 4.7 (1.1) | -0.02 (-0.21, 0.17) |
| Communication comfort | 277 | 5.7 (1.0) | 363 | 5.6 (1.0) | -0.05 (-0.21, 0.12) |
| Rapport | 275 | 5.6 (1.0) | 361 | 5.6 (1.0) | -0.05 (-0.22, 0.12) |
| Compliance intent | 274 | 5.4 (1.1) | 360 | 5.3 (1.0) | -0.15 (-0.32, 0.03) |
| **Process measures** |  |  |  |  |  |
| **Perceptions of empathy (CARE)** | 275 | 42.5 (9.5) | 361 | 41.2 (9.8) | -1.57 (-3.34, 0.20) |
| **Perceptions of practitioner optimism** | 182 | 5.5 (1.0) | 239 | 5.3 (1.1) | -0.16 (-0.37, 0.06) |
| **Treatment expectation (TEX-Q)** | 174 | 7.1 (1.7) | 226 | 7.1 (1.6) | 0.01 (-0.32, 0.34) |
| Treatment benefit | 178 | 6.5 (2.6) | 234 | 6.4 (2.6) | -0.09 (-0.64, 0.46) |
| Positive impact | 179 | 5.6 (3.1) | 232 | 5.8 (3.1) | 0.17 (-0.43, 0.78) |
| Adverse events | 179 | 8.0 (2.2) | 236 | 7.9 (2.3) | -0.14 (-0.58, 0.30) |
| Negative impact | 177 | 8.6 (2.1) | 235 | 8.5 (2.1) | -0.13 (-0.57, 0.31) |
| Process | 177 | 7.2 (2.0) | 235 | 7.3 (2.2) | 0.03 (-0.38, 0.44) |
| Behaviour centre | 178 | 7.0 (2.6) | 237 | 7.0 (2.7) | 0.03 (-0.48, 0.54) |
| **Patient doctor depth of relationship** | 267 | 17.3 (8.1) | 349 | 18.2 (8.3) | 0.41 (-1.45, 2.28) |
| **Anxiety (HADS-A)** | 272 | 6.5 (4.6) | 354 | 6.5 (4.7) | -0.17 (-1.13, 0.79) |
| **Depression (HADS-D)** | 272 | 4.6 (4.1) | 353 | 4.6 (4.3) | -0.03 (-0.85, 0.79) |

*Adjusted for stratification factors (practice size, practice deprivation); Patient Enablement Index is on a scale of 1 (least enablement) to 7 (most enablement)

Patient global impression (PGI) of symptom severity is scored from 1 (none) to 7 (extremely severe)

PGI of symptom change is scored from 1 (very much improved) to 7 (very much worse)

Pain interference ranges from 0 to 10, with higher scores indicating greater interference

Patient satisfaction (MISS) ranges from 1 to 7, with higher scores indicating greater satisfaction

Perceptions of practitioner empathy (CARE) ranges from 10 to 50 with higher scores indicating greater perceived empathy

Perceptions of practitioner optimism ranges 1 (extremely pessimistic) to 7 (extremely optimistic)

Treatment expectations (TEX-Q) ranges from 0 to 10, with higher values indicating more positive treatment expectations

HADS Anxiety and depression subscales range from 0 to 21, with the following interpretation: 0-7 (Normal), 8-10 (Mild), 11-15 (Moderate), 16-21 (Severe)

Patient doctor depth of relationship ranges from 0 (no relationship) to 32 (very strong relationship)

### Supplementary Table 4. Subgroup analyses for MSK and All-comer participants – BPI average pain

| BPI average pain | N | BPI average pain  Mean difference (97.5% CI) | Adjusted* interaction term – mean difference (97.5% CI) |
| --- | --- | --- | --- |
| Gender |  |  |  |
| Male | 315 | -0.31 (-0.69, 0.08) | REF |
| Female | 716 | 0.20 (-0.05, 0.45) | 0.48 (0.03, 0.94) |
| Age group |  |  |  |
| <45 | 174 | 0.74 (0.12, 1.34) | REF |
| 45- <65 | 437 | -0.06 (-0.39, 0.26) | -0.72 (-1.34, -0.11) |
| 65+ | 422 | -0.08 (-0.45, 0.29) | -0.72 (-1.34, -0.11) |
| IMD group |  |  |  |
| 1 - 5 (higher deprivation) | 347 | 0.31 (-0.06, 0.68) | REF |
| 6 - 10 (lower deprivation) | 652 | -0.13 (-0.41, 0.14) | -0.41 (-0.86, 0.04) |
| Consultation type |  |  |  |
| In person | 828 | -0.02 (-0.26, 0.21) | REF |
| Telephone | 206 | 0.36 (-0.09, 0.81) | 0.26 (-0.26, 0.79) |
| HCP type |  |  |  |
| Doctor | 540 | -0.02 (-0.33, 0.30) |  |
| Nurse | 30 | -0.05 (-1.29 1.19) | -0.13 (-1.35, 1.10) |
| Physiotherapist | 275 | -0.03 (-0.44, 0.39) | 0.01 (-0.49, 0.51) |
| Baseline pain |  |  |  |
| BPI average 0 - 3 | 192 | -0.12 (-0.58, 0.35) | REF |
| BPI average 4 - 7 | 675 | 0.03 (-0.22, 0.29) | 0.13 (-0.41, 0.67) |
| BPI average 8 - 10 | 167 | 0.20 (-0.36, 0.75) | 0.33 (-0.39, 1.03) |
| Consultation reason |  |  |  |
| Other | 923 | 0.06 (-0.16, 0.28) | REF |
| General | 96 | -0.24 (-0.96, 0.48) | 0.13 (-0.37, 0.62) |
| Mental | 12 | -0.37 (-1.73, 0.99) | -0.42 (-1.62, 0.79) |

*Adjusted for stratification factors (practice size and practice deprivation score).

### Supplementary Table 5. Subgroup analyses for MSK and All-comer participants – Patient enablement index

| Patient enablement index | N | PEI  Mean difference (97.5% CI) | Adjusted* interaction term – mean difference (97.5% CI) |
| --- | --- | --- | --- |
| Gender |  |  |  |
| Male | 460 | 0.14 (-0.09, 0.37) | REF |
| Female | 938 | 0.02 (-0.18, 0.22) | -0.11 (-0.39, 0.18) |
| Age group |  |  |  |
| <45 | 246 | -0.09 (-0.46, 0.29) | REF |
| 45- <65 | 573 | 0.06 (-0.15, 0.28) | 0.13 (-0.26, 0.51) |
| 65+ | 582 | 0.12 (-0.07, 0.32) | 0.18 (-0.21, 0.56) |
| IMD group |  |  |  |
| 1 - 5 (higher deprivation) | 465 | -0.01 (-0.30, 0.29) | REF |
| 6 - 10 (lower deprivation) | 890 | 0.08 (-0.08, 0.25) | 0.09 (-0.21, 0.39) |
| Consultation modality |  |  |  |
| In person | 1064 | 0.11 (-0.09, 0.31) | REF |
| Telephone | 336 | -0.07 (-0.36, 0.22) | -0.13 (-0.46, 0.20) |
| HCP type |  |  |  |
| Doctor | 812 | 0.11 (-0.08, 0.30) | REF |
| Nurse | 67 | 0.56 (-0.05, 1.17) | 0.42 (-0.22, 1.07) |
| Physiotherapist | 273 | 0.10 (-0.28, 0.48) | -0.01 (-0.38, 0.37) |
| Baseline pain |  |  |  |
| BPI average 0 - 3 | 185 | -0.12 (-0.58, 0.35) | REF |
| BPI average 4 - 7 | 658 | 0.03 (-0.22, 0.29) | 0.13 (-0.41, 0.67) |
| BPI average 8 - 10 | 161 | 0.20 (-0.36, 0.75) | 0.33 (-0.39, 1.03) |
| Consultation reason |  |  |  |
| Other | 1159 | 0.06 (-0.12, 0.24) | REF |
| General | 195 | 0.06 (-0.30, 0.43) | -0.02 (-0.41, 0.37) |
| Mental | 48 | -0.01 (-0.73, 0.70) | -0.06 (-0.81, 0.69) |

*Adjusted for stratification factors (practice size and practice deprivation score)

### Supplementary Table 6. Serious adverse events by trial arm for musculoskeletal and All-comers groups

|  | Usual care  n (%) | Intervention  n (%) | Adjusted* odds ratio (95% CI) |
| --- | --- | --- | --- |
| **Musculoskeletal pain group** |  |  |  |
| Participants experiencing at least one SAE, n(%) | 15/439 (3.4) | 12/367 (3.3) | 0.93 (0.43, 2.03) |
| Death | 0 | 1 |  |
| Admission to hospital | 7 | 7 |  |
| Life threatening event | 3 | 3 |  |
| Other medical event | 5 | 1 |  |
| **All-comers group** |  |  |  |
| Participants experiencing at least one SAE, n(%) | 16/386 (4.2) | 17/490 (3.5) | 0.84 (0.42, 1.68) |
| Death | 0 | 0 |  |
| Admission to hospital | 10 | 9 |  |
| Life threatening event | 4 | 4 |  |
| Other medical event | 2 | 4 |  |

Note. All participants who experienced an SAE had one SAE, except 5 participants who had two SAEs (5 additional SAEs: 4 hospital admissions, 1 life threatening event)

### Supplementary Table 7. Unit costs for health economic analysis

| **Resource** | **Unit cost (£)** |
| --- | --- |
| Accident and emergency | 268 |
| Outpatient (in-person) | 171 |
| Outpatient (virtual) | 140 |
| Day case | 672 |
| Inpatient (per day) | 414 |
| GP (at the practice) | 49 |
| GP (at home) | 116 |
| GP (virtual) | 35 |
| Other healthcare professional (at the practice) | 18 |
| Other healthcare professional (at home) | 45 |
| Other healthcare professional (virtual) | 5 |
| Prescribed medications | By medication |
| Time off work (per hour) | 20 |

### Supplementary Table 8. Mean EQ-5D-5L, ICECAP-A, QALYs and capability by intervention allocation for musculoskeletal and All comers groups

|  |  | **Intervention** | | | | **Usual care** | | | |
| --- | --- | --- | --- | --- | --- | --- | --- | --- | --- |
|  |  | **n** | **(%)** | **Mean** | **(SD)** | **n** | **(%)** | **Mean** | **(SD)** |
| **Musculoskeletal group** | | | | | | | | | |
| EQ-5D-5L | |  |  |  |  |  |  |  |  |
|  | Baseline | 367 | (100) | 0.493 | (0.266) | 439 | (100) | 0.502 | (0.256) |
|  | 1 month | 236 | (64) | 0.564 | (0.284) | 320 | (73) | 0.538 | (0.269) |
|  | 6 months | 226 | (62) | 0.564 | (0.291) | 279 | (64) | 0.578 | (0.268) |
| QALYs | | 186 | (51) | 0.286 | (0.128) | 254 | (58) | 0.280 | (0.116) |
| ICECAP-A | | | | | | | | | |
|  | Baseline | 365 | (99) | 0.771 | (0.206) | 437 | (100) | 0.781 | (0.187) |
|  | 1 month | 232 | (63) | 0.771 | (0.207) | 318 | (72) | 0.772 | (0.193) |
|  | 6 months | 222 | (60) | 0.769 | (0.218) | 276 | (63) | 0.790 | (0.194) |
| Years of full capability equivalent | | 179 | (49) | 0.390 | (0.099) | 250 | (57) | 0.394 | (0.088) |
| **All comers group** | | | | | | | | | |
| EQ-5D-5L | |  |  |  |  |  |  |  |  |
|  | Baseline | 490 | (100) | 0.706 | (0.247) | 386 | (100) | 0.732 | (0.201) |
|  | 1 month | 341 | (70) | 0.729 | (0.252) | 265 | (69) | 0.754 | (0.223) |
|  | 6 months | 298 | (61) | 0.730 | (0.260) | 229 | (59) | 0.733 | (0.239) |
| QALYs | | 274 | (56) | 0.366 | (0.118) | 207 | (54) | 0.375 | (0.105) |
| ICECAP-A | | | | | | | | | |
|  | Baseline | 487 | (99) | 0.830 | (0.188) | 384 | (99) | 0.847 | (0.162) |
|  | 1 month | 340 | (69) | 0.836 | (0.189) | 262 | (68) | 0.845 | (0.160) |
|  | 6 months | 296 | (60) | 0.833 | (0.188) | 229 | (59) | 0.858 | (0.166) |
| Years of full capability equivalent | | 270 | (55) | 0.419 | (0.088) | 206 | (53) | 0.430 | (0.075) |

### Supplementary Table 9. Mean resource use and costs by intervention allocation for the musculoskeletal group

|  | **Intervention (n=367)** | | | | | **Control (n=439)** | | | | |
| --- | --- | --- | --- | --- | --- | --- | --- | --- | --- | --- |
|  | **n** | **(%)** | **Number of contacts** | **Mean cost (£) (SD)** | | **n** | **(%)** | **Number of contacts** | **Mean cost (£) (SD)** | |
| Accident and emergency | 187 | (51) | 0.32 | 81 | (261) | 240 | (55) | 0.30 | 78 | (202) |
| Outpatient (in-person) | 185 | (50) | 2.13 | 363 | (475) | 242 | (55) | 2.55 | 435 | (704) |
| Outpatient (virtual) | 184 | (50) | 0.75 | 105 | (176) | 241 | (55) | 0.82 | 115 | (188) |
| Day case | 187 | (51) | 0.23 | 155 | (618) | 243 | (55) | 0.19 | 130 | (391) |
| Inpatient (per day) | 186 | (51) | 0.07 | 62 | (481) | 243 | (55) | 0.11 | 115 | (488) |
| GP (GP surgery, health or walk-in centre) | 187 | (51) | 2.77 | 137 | (126) | 242 | (55) | 2.68 | 132 | (127) |
| GP (home) | 188 | (51) | 0.16 | 19 | (93) | 242 | (55) | 0.09 | 11 | (45) |
| GP (virtual) | 186 | (51) | 1.62 | 60 | (77) | 242 | (55) | 1.61 | 60 | (75) |
| OHCP* (GP surgery, health or walk-in centre) | 186 | (51) | 2.34 | 42 | (43) | 242 | (55) | 2.39 | 43 | (43) |
| OHCP* (at home) | 188 | (51) | 0.23 | 1 | (3) | 241 | (55) | 0.28 | 1 | (5) |
| OHCP* (virtual) | 186 | (51) | 0.49 | 22 | (41) | 243 | (55) | 0.55 | 25 | (53) |
| Prescribed medications | 184 | (50) |  | 149 | (351) | 241 | (55) |  | 148 | (404) |
| Total NHS cost | 179 | (49) |  | 1101 | (1091) | 233 | (53) |  | 1278 | (1504) |
| Travel for healthcare contacts | 171 | (47) |  | 24 | (63) | 223 | (51) |  | 32 | (92) |
| Private healthcare | 182 | (50) |  | 293 | (1809) | 237 | (54) |  | 230 | (1308) |
| Over-the-counter medications | 138 | (38) |  | 26 | (44) | 191 | (44) |  | 22 | (29) |
| Missed work due to health problems | 181 | (49) |  | 535 | (2304) | 240 | (55) |  | 764 | (2612) |
| Total societal cost | 130 | (35) |  | 1866 | (3249) | 169 | (38) |  | 2318 | (3975) |
| *OHCP: Other healthcare professional |  |  |  |  |  |  |  |  |  |  |

### Supplementary Table 10. Cost-effectiveness analysis for musculoskeletal and All comers groups

|  | **Intervention** | | | **Usual care** | | | **Adjusted difference**  **(95% CI)** | |
| --- | --- | --- | --- | --- | --- | --- | --- | --- |
|  | **n** | **Adjusted mean (SE)** | | **n** | **Adjusted mean (SE)** | |  |  |
| **Musculoskeletal group** | | | | | | | | |
| Health care cost (£) | 367 | 1329 | (103) | 439 | 1433 | (107) | -104 | (-393 to 186) |
| Years of full capability equivalent | 367 | 0.386 | (0.005) | 439 | 0.382 | (0.004) | 0.003 | (-0.011 to 0.017) |
| ICER | Intervention dominates usual care | | | | | | | |
| **All comers group** | | | | | | | | |
| Health care cost (£) | 490 | 1247 | (109) | 386 | 1271 | (149) | -23 | (-395 to 348) |
| Years of full capability equivalent | 490 | 0.416 | (0.003) | 386 | 0.417 | (0.003) | -0.001 | (-0.011 to 0.009) |
| ICER | Intervention is less costly but has fewer years of full capability equivalent. £19223 saved per year of full capability equivalent lost | | | | | | | |
| Adjusted for baseline ICECAP-A, baseline BPI (MSK group only), practice deprivation (high/low), practice size (large/small) | | | | | | | | |

### Supplementary Table 11. Cost-consequences based on all available data for musculoskeletal and All comers groups

|  |  | **Intervention** | | | **Control** | | | **Difference  (95% CI)** | | **Adjusted difference  (95% CI)** | |
| --- | --- | --- | --- | --- | --- | --- | --- | --- | --- | --- | --- |
|  |  | **n** | **Mean (SD)** | | **n** | **Mean (SD)** | |  |  |  |  |
| **Musculoskeletal group** | | | | | | | | | | | |
| Cost (£) | |  |  |  |  |  |  |  |  |  |  |
|  | EMPathicO intervention | 367 | 0.42 | (0) |  |  |  |  |  |  |  |
|  | NHS costs | 179 | 1102 | (1091) | 233 | 1278 | (1504) | -177 | (-85 to 439) |  |  |
|  | Travel for healthcare contacts | 171 | 24 | (63) | 223 | 32 | (92) | -9 | (-7 to 25) |  |  |
|  | Private healthcare | 182 | 293 | (1809) | 237 | 230 | (1308) | 63 | (-363 to 236) |  |  |
|  | Over-the-counter medications | 138 | 26 | (44) | 191 | 22 | (29) | 4 | (-12 to 4) |  |  |
|  | Missed work due to health problems | 181 | 535 | (2304) | 240 | 764 | (2612) | -229 | (-252 to 709) |  |  |
| Outcomes | |  |  |  |  |  |  |  |  |  |  |
|  | Pain intensity (Repeated measures) | 388 |  |  | 318 |  |  |  |  | 0.06 | 97.5% CI (-0.19 to 0.31) |
|  | Patient enablement (repeated measures) | 380 |  |  | 309 |  |  |  |  | 0.17 | 97.5% CI (-0.05 to 0.40) |
|  | Quality-adjusted life years | 186 | 0.286 | (0.128) | 254 | 0.280 | (0.116) |  |  | 0.011 | (-0.004 to 0.026)* |
|  | Years of full capability equivalent | 179 | 0.390 | (0.099) | 250 | 0.394 | (0.088) |  |  | 0.001 | (-0.009 to 0.011)* |
| **All comers group** | | | | | | | | | | | |
| Cost (£) | |  |  |  |  |  |  |  |  |  |  |
|  | EMPathicO intervention | 367 | 0.42 | (0) |  |  |  |  |  |  |  |
|  | NHS costs | 251 | 1132 | (1907) | 196 | 1313 | (3363) | -180 | (-315 to 676) |  |  |
|  | Travel for healthcare contacts | 229 | 22 | (79) | 192 | 17 | (57) | 4 | (-17 to 9) |  |  |
|  | Private healthcare | 247 | 146 | (951) | 197 | 77 | (389) | 69 | (-211 to 72) |  |  |
|  | Over-the-counter medications | 225 | 16 | (25) | 168 | 13 | (20) | 3 | (-8 to 1) |  |  |
|  | Missed work due to health problems | 251 | 530 | (2253) | 202 | 231 | (931) | 299 | (-631 to 34) |  |  |
| Outcomes | |  |  |  |  |  |  |  |  |  |  |
|  | Patient enablement (repeated measures) | 316 |  |  | 397 |  |  |  |  | -0.12 | (-0.32 to 0.07) |
|  | Quality-adjusted life years | 274 | 0.366 | (0.118) | 207 | 0.375 | (0.105) |  |  | 0.003 | (-0.007 to 0.013)* |
|  | Years of full capability equivalent | 270 | 0.419 | (0.088) | 206 | 0.430 | (0.075) |  |  | 0.005 | (-0.004 to 0.016)* |
| *Adjusted for baseline EQ-5D-5L/ICECAP-A, baseline BPI (MSK group only), practice deprivation (high/low), practice size (large/small) | | | | | | | | | | | |

### Supplementary Table 12. Incremental costs, QALYs and net benefit from an NHS perspective, by intervention allocation for All comers groups

|  |  | **Intervention** | | | **Usual care** | | |
| --- | --- | --- | --- | --- | --- | --- | --- |
|  |  | **n** | **Mean** | **(SD/SE)** | **n** | **Mean** | **(SD/SE)** |
| EMPathicO cost | | 367 | 0.42 | (0) |  |  |  |
| Primary care cost (£) | | 256 | 243 | (236) | 197 | 234 | (234) |
| Secondary care cost (£) | | 259 | 806 | (1825) | 202 | 927 | (3225) |
| Prescribed medication cost (£) | | 258 | 118 | (166) | 204 | 154 | (374) |
| Adjusted health care cost (£)* | | 490 | 1247 | (109) | 386 | 1271 | (149) |
| Adjusted difference in health care cost (£)* | | | | -23 | (-395 to 348) | |  |
| EQ-5D-5L | |  |  |  |  |  |  |
|  | Baseline | 490 | 0.706 | (0.247) | 386 | 0.732 | (0.201) |
|  | 1 month | 341 | 0.729 | (0.252) | 265 | 0.754 | (0.223) |
|  | 6 months | 298 | 0.730 | (0.260) | 229 | 0.733 | (0.239) |
| Adjusted QALYs | | 490 | 0.364 | (0.003) | 386 | 0.361 | (0.003) |
| Adjusted difference in QALYs* | | | | 0.004 | (-0.005 to 0.013) | | |
| INMB at £20,000 per QALY (95% CI) (£)* | | | | 134 | (-298 to 565) P = 0.54 | | |
| INMB at £30,000 per QALY (95% CI) (£)* | | | | 170 | (-313 to 652) P = 0.49 | | |
| Standard deviation (SD) presented for complete case data. Standard error (SE) presented for imputed data. INMB: incremental net monetary benefit | | | | | | | |
| *Generalised linear models after multiple imputation, adjusted for baseline EQ-5D-5L (QALY/iNMB only), baseline BPI (MSK group only), practice deprivation (high/low), practice size (large/small) | | | | | | | |

### Supplementary Table 13. Mean resource use and costs by intervention allocation for the All comers group

|  | **Intervention (n=490)** | | | | | **Control (n=386)** | | | | |
| --- | --- | --- | --- | --- | --- | --- | --- | --- | --- | --- |
|  | **n** | **(%)** | **Number of contacts** | **Mean cost (£) (SD)** | | **n** | **(%)** | **Number of contacts** | **Mean cost (£) (SD)** | |
| Accident and emergency | 262 | (53) | 0.30 | 78 | (227) | 204 | (53) | 0.26 | 67 | (220) |
| Outpatient (in-person) | 262 | (53) | 1.52 | 258 | (541) | 204 | (53) | 1.58 | 270 | (421) |
| Outpatient (virtual) | 262 | (53) | 0.66 | 92 | (175) | 205 | (53) | 0.77 | 107 | (190) |
| Day case | 262 | (53) | 0.18 | 123 | (370) | 203 | (53) | 0.15 | 103 | (342) |
| Inpatient | 262 | (53) | 0.12 | 249 | (1372) | 204 | (53) | 0.10 | 377 | (2977) |
| GP (GP surgery, health or walk-in centre) | 260 | (53) | 2.40 | 118 | (126) | 201 | (52) | 2.55 | 126 | (135) |
| GP (home) | 262 | (53) | 0.09 | 11 | (50) | 204 | (53) | 0.07 | 8 | (61) |
| GP (virtual) | 260 | (53) | 1.51 | 56 | (70) | 202 | (52) | 1.32 | 49 | (70) |
| OHCP* (GP surgery, health or walk-in centre) | 258 | (53) | 2.03 | 36 | (60) | 203 | (53) | 2.09 | 37 | (38) |
| OHCP* (at home) | 261 | (53) | 0.23 | 1 | (6) | 204 | (53) | 0.20 | 1 | (4) |
| OHCP* (virtual) | 260 | (53) | 0.56 | 25 | (51) | 202 | (52) | 0.52 | 23 | (49) |
| Prescribed medications | 258 | (53) |  | 118 | (166) | 204 |  |  | 154 | (374) |
| Total NHS cost | 251 | (51) |  | 1132 | (1907) | 196 | (51) |  | 1313 | (3363) |
| Travel for healthcare contacts | 229 | (47) |  | 22 | (79) | 192 | (50) |  | 17 | (57) |
| Private healthcare | 247 | (50) |  | 146 | (951) | 197 | (51) |  | 77 | (389) |
| Over-the-counter medications | 225 | (46) |  | 16 | (25) | 168 | (44) |  | 13 | (20) |
| Missed work due to health problems | 251 | (51) |  | 530 | (2253) | 202 | (52) |  | 231 | (931) |
| Total societal cost | 194 | (40) |  | 1629 | (3087) | 152 | (39) |  | 1689 | (3981) |
| *OHCP: Other healthcare professional |  |  |  |  |  |  |  |  |  |  |

### Supplementary Table 14 Secondary and sensitivity economic analyses, by intervention allocation for All-comers groups

|  | **Intervention (n=367)** | | | **Usual care (n=439)** | | | **Adjusted difference**  **(95% CI)** | | **iNMB (95% CI) (£)** | |
| --- | --- | --- | --- | --- | --- | --- | --- | --- | --- | --- |
|  | **n** | **Adjusted mean (SE)** | | **n** | **Adjusted mean (SE)** | |  |  | **£20,000 per QALY** | **30,000 per QALY** |
| Secondary analysis: Societal (NHS, personal expenses and productivity) perspective | | | | | | | | | | |
| Societal cost (£) | 490 | 2007 | (210) | 386 | 1745 | (196) | 262 | (-296 to 820) | 87 | 123 |
| QALYs | 490 | 0.365 | (0.003) | 386 | 0.361 | (0.003) | 0.004 | (-0.005 to 0.013) | (-446 to 621) | (-453 to 699) |
| Sensitivity analysis: NHS perspective including complete case data only | | | | | | | | | | |
| Health care cost | 251 | 1160 | (141) | 196 | 1276 | (204) | -116 | (-689 to 242) | 292 | 302 |
| QALYs | 274 | 0.371 | (0.004) | 207 | 0.368 | (0.003) | 0.003 | (-0.007 to 0.013) | (-268 to 1059) | (-329 to 1118) |
| Sensitivity analysis (post-hoc): NHS primary care perspective | | | | | | | | | | |
| Primary care cost (£) | 490 | 370 | (21) | 386 | 425 | (28) | -55 | (-127 to 17) | 200 | 236 |
| QALYs | 490 | 0.365 | (0.003) | 386 | 0.361 | (0.003) | 0.004 | (-0.005 to 0.012) | (-140 to 539) | (-159 to 631) |
| Standard deviation (SD) presented for complete case data. Standard error (SE) presented for imputed data. iNMB: incremental net monetary benefit | | | | | | | | | | |
| Generalised linear models after multiple imputation, adjusted for baseline EQ-5D-5L (QALY/iNMB only), baseline BPI (MSK group only), practice deprivation (high/low), practice size (large/small) | | | | | | | | | | |

### Supplementary Figure 1 Cost-effectiveness acceptability curve for All comers group

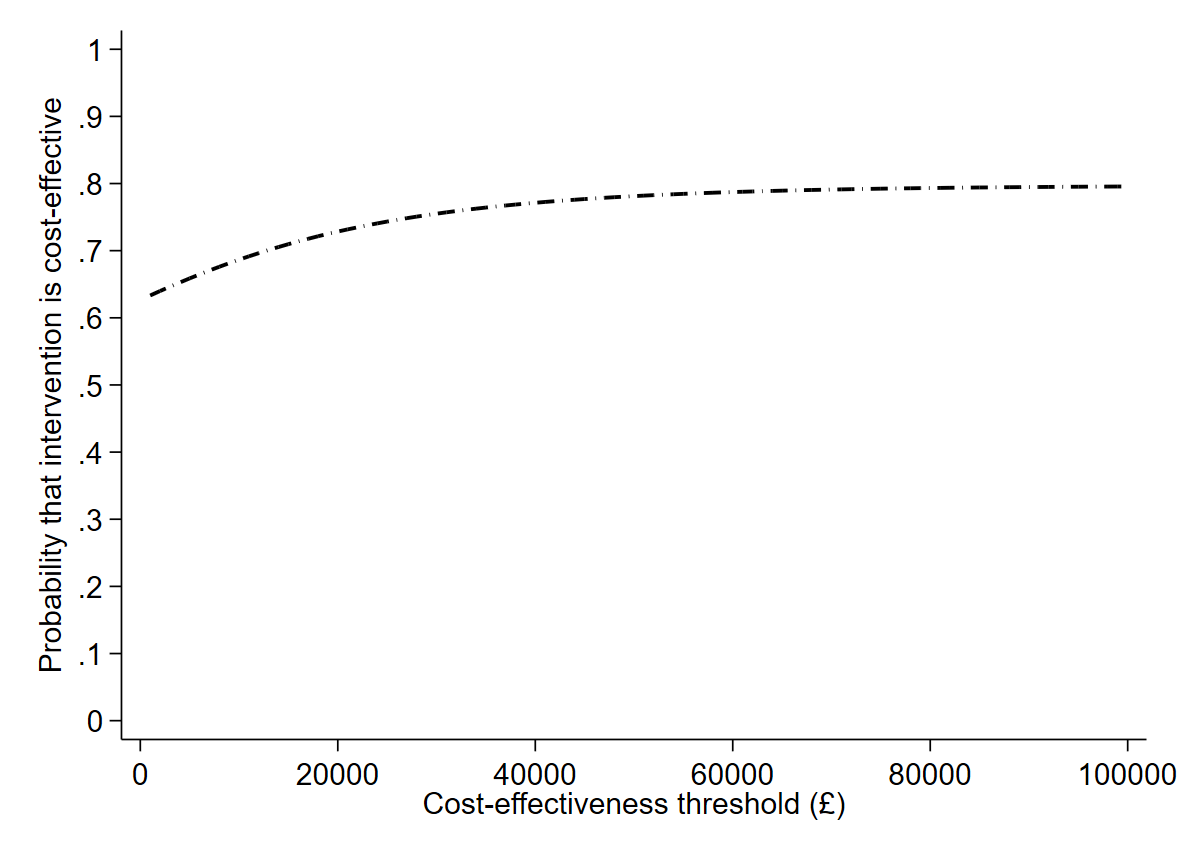
