## Supplementary material for "Clinical and cost-effectiveness of communication skills e-learning for primary care practitioners on patients’ musculoskeletal pain and enablement: the Talking in Primary care (TIP) cluster-randomised controlled trial": Consort cluster trials checklist

### CONSORT 2010 checklist cluster randomised trial extension

| **Section/topic and item No** | **Standard checklist item** | **Extension for cluster designs** | **Page No*** |
| --- | --- | --- | --- |
| **Title and abstract** | | | |
| 1a | Identification as a randomised trial in the title | Identification as a cluster randomised trial in the title | 1 |
| 1b | Structured summary of trial design, methods, results, and conclusions (for specific guidance see CONSORT for abstracts)^11 12^ | See table 2 | 4-5 |
| **Introduction** | | | |
| Background and objectives: |  |  |  |
| 2a | Scientific background and explanation of rationale | Rationale for using a cluster design | 6 |
| 2b | Specific objectives or hypotheses | Whether objectives pertain to the cluster level, the individual participant level, or both | 7 |
| **Methods** | | | |
| Trial design: |  |  |  |
| 3a | Description of trial design (such as parallel, factorial) including allocation ratio | Definition of cluster and description of how the design features apply to the clusters | 7 |
| 3b | Important changes to methods after trial commencement (such as eligibility criteria), with reasons |  | 7 |
| Participants: |  |  |  |
| 4a | Eligibility criteria for participants | Eligibility criteria for clusters | 8-9 |
| 4b | Settings and locations where the data were collected |  | 8-9 |
| Interventions: |  |  |  |
| 5 | The interventions for each group with sufficient details to allow replication, including how and when they were actually administered | Whether interventions pertain to the cluster level, the individual participant level, or both | 10 |
| Outcomes: |  |  |  |
| 6a | Completely defined prespecified primary and secondary outcome measures, including how and when they were assessed | Whether outcome measures pertain to the cluster level, the individual participant level, or both | 11-14 |
| 6b | Any changes to trial outcomes after the trial commenced, with reasons |  | N/A |
| Sample size: |  |  |  |
| 7a | How sample size was determined | Method of calculation, number of clusters(s) (and whether equal or unequal cluster sizes are assumed), cluster size, a coefficient of intracluster correlation (ICC or *k*), and an indication of its uncertainty | 14-15 |
| 7b | When applicable, explanation of any interim analyses and stopping guidelines |  | N/A |
| **Randomisation** | | | |
| Sequence generation: |  |  |  |
| 8a | Method used to generate the random allocation sequence |  | 15 |
| 8b | Type of randomisation; details of any restriction (such as blocking and block size) | Details of stratification or matching if used | 15 |
| Allocation concealment mechanism: |  |  |  |
| 9 | Mechanism used to implement the random allocation sequence (such as sequentially numbered containers), describing any steps taken to conceal the sequence until interventions were assigned | Specification that allocation was based on clusters rather than individuals and whether allocation concealment (if any) was at the cluster level, the individual participant level, or both | 15 |
| Implementation: |  |  |  |
| 10 | Who generated the random allocation sequence, who enrolled participants, and who assigned participants to interventions | Replaced by 10a, 10b, and 10c |  |
| 10a |  | Who generated the random allocation sequence, who enrolled clusters, and who assigned clusters to interventions | 15 |
| 10b |  | Mechanism by which individual participants were included in clusters for the purposes of the trial (such as complete enumeration, random sampling) | 9 |
| 10c |  | From whom consent was sought (representatives of the cluster, or individual cluster members, or both) and whether consent was sought before or after randomisation | 8,9 |
| Blinding: |  |  |  |
| 11a | If done, who was blinded after assignment to interventions (for example, participants, care providers, those assessing outcomes) and how |  | 15 |
| 11b | If relevant, description of the similarity of interventions |  | N/A |
| Statistical methods: |  |  |  |
| 12a | Statistical methods used to compare groups for primary and secondary outcomes | How clustering was taken into account | 16 |
| 12b | Methods for additional analyses, such as subgroup analyses and adjusted analyses |  | 16,17 |
| **Results** | | | |
| Participant flow (a diagram is strongly recommended): |  |  |  |
| 13a | For each group, the numbers of participants who were randomly assigned, received intended treatment, and were analysed for the primary outcome | For each group, the numbers of clusters that were randomly assigned, received intended treatment, and were analysed for the primary outcome | 18,19, Fig1, Fig 2 |
| 13b | For each group, losses and exclusions after randomisation, together with reasons | For each group, losses and exclusions for both clusters and individual cluster members | 18,19, Fig1, Fig 2 |
| Recruitment: |  |  |  |
| 14a | Dates defining the periods of recruitment and follow-up |  | 18,19 |
| 14b | Why the trial ended or was stopped |  | 19 |
| Baseline data: |  |  |  |
| 15 | A table showing baseline demographic and clinical characteristics for each group | Baseline characteristics for the individual and cluster levels as applicable for each group | Table 1, Table 2, Supp Table 1 |
| Numbers analysed: |  |  |  |
| 16 | For each group, number of participants (denominator) included in each analysis and whether the analysis was by original assigned groups | For each group, number of clusters included in each analysis | Table 1, Table 2, Supp Table 1 |
| Outcomes and estimation: |  |  |  |
| 17a | For each primary and secondary outcome, results for each group, and the estimated effect size and its precision (such as 95% confidence interval) | Results at the individual or cluster level as applicable and a coefficient of intracluster correlation (ICC or *k*) for each primary outcome | Table 3, Supp Table 3 |
| 17b | For binary outcomes, presentation of both absolute and relative effect sizes is recommended |  | N/A |
| Ancillary analyses: |  |  |  |
| 18 | Results of any other analyses performed, including subgroup analyses and adjusted analyses, distinguishing prespecified from exploratory |  | Tables 4,5,6 Supp Tables 4,5, 7-14 |
| Harms: |  |  |  |
| 19 | All important harms or unintended effects in each group (for specific guidance see CONSORT for harms106) |  | 26,Supp Table 6 |
| **Discussion** | | | |
| Limitations: |  |  |  |
| 20 | Trial limitations, addressing sources of potential bias, imprecision, and, if relevant, multiplicity of analyses |  | 30 |
| Generalisability: |  |  |  |
| 21 | Generalisability (external validity, applicability) of the trial findings | Generalisability to clusters and/or individual participants (as relevant) | 30 |
| Interpretation: |  |  |  |
| 22 | Interpretation consistent with results, balancing benefits and harms, and considering other relevant evidence |  | 31,32 |
| **Other information** | | | |
| Registration: |  |  |  |
| 23 | Registration number and name of trial registry |  | 5,33 |
| Protocol: |  |  |  |
| 24 | Where the full trial protocol can be accessed, if available |  | 33 |
| Funding: |  |  |  |
| 25 | Sources of funding and other support (such as supply of drugs), role of funders |  | 33,34 |

*Page numbers optional depending on journal requirements.
