## Supplementary material for "Clinical and cost-effectiveness of communication skills e-learning for primary care practitioners on patients’ musculoskeletal pain and enablement: the Talking in Primary care (TIP) cluster-randomised controlled trial": Consort abstracts checklist

Extension of CONSORT for abstracts to reports of cluster randomised trials

| **Item** | **Standard checklist item** | **Extension for cluster trials** | **Page** |
| --- | --- | --- | --- |
| Title | Identification of study as randomised | Identification of study as cluster randomised | 1 |
| Trial design | Description of the trial design (for example, parallel, cluster, non-inferiority) |  | 4 |
| Methods: |  |  |  |
| Participants | Eligibility criteria for participants and the settings where the data were collected | Eligibility criteria for clusters | 4 |
| Interventions | Interventions intended for each group |  | 4 |
| Objective | Specific objective or hypothesis | Whether objective or hypothesis pertains to the cluster level, the individual participant level, or both | 4 |
| Outcome | Clearly defined primary outcome for this report | Whether the primary outcome pertains to the cluster level, the individual participant level or both | 4 |
| Randomisation | How participants were allocated to interventions | How clusters were allocated to interventions | 4 |
| Blinding (masking) | Whether or not participants, care givers, and those assessing the outcomes were blinded to group assignment |  | 4 |
| Results: |  |  |  |
| Numbers randomised | Number of participants randomised to each group | Number of clusters randomised to each group | 5 |
| Recruitment | Trial status* |  | 5 |
| Numbers analysed | Number of participants analysed in each group | Number of clusters analysed in each group | 5 |
| Outcome | For the primary outcome, a result for each group and the estimated effect size and its precision | Results at the cluster or individual level as applicable for each primary outcome | 5 |
| Harms | Important adverse events or side effects |  | 5 |
| Conclusions | General interpretation of the results |  | 5 |
| Trial registration | Registration number and name of trial register |  | 5 |
| Funding | Source of funding |  | 5 |

*Relevant to conference abstracts.
