## Supplementary material for "Clinical and cost-effectiveness of communication skills e-learning for primary care practitioners on patients’ musculoskeletal pain and enablement: the Talking in Primary care (TIP) cluster-randomised controlled trial": TIDieR statement intervention description

EMPathicO (Communicating clinical EMPathy and realistic Optimism in primary care consultations)

TIDieR

### EMPathicO (Communicating clinical EMPathy and realistic Optimism in primary care consultations)

**Why:**

#### Rationale for elements essential to the intervention to change practitioner behaviour

1. Evidence-based information about the consequences of communicating clinical empathy and realistic optimism and verbal persuasion that communication skills can facilitate efficient consultations, may enhance primary care practitioners' expectancies that it is feasible to communicate clinical empathy and realistic optimism in primary care consultations and that doing so can enhance patient outcomes.
2. Acknowledging barriers to enacting communication skills and supporting practitioners' problem-solving may enhance practitioner self-efficacy to communicate clinical empathy and realistic optimism in consultations.
3. Providing instruction and (video) demonstration of how to communicate clinical empathy and realistic optimism in consultations, and user testimonials may enhance practitioner skills in communicating clinical empathy and realistic optimism in consultations.
4. Enabling practitioners to monitor their communication behaviour and supporting them to reflect on their communication behaviours, to set goals and action plan changing their communication behaviours may strengthen practitioner behavioural intentions to communicate clinical empathy and realistic optimism in consultations.
5. Enhancing practitioners' expectancies, self-efficacy, skills, and intentions about communicating clinical empathy and realistic optimism may enhance their actual communication of clinical empathy and realistic optimism in consultations.

#### Rationale for practitioner behaviour changing patient

#### outcomes

1. Enhancing practitioners' expressions of clinical empathy may increase patients' perceptions of empathy and decrease patient anxiety.
2. Enhancing practitioners' expressions of realistic empathy may increase patients' perceptions of practitioner optimism and decrease patient anxiety.
3. Increasing patients' perceptions of practitioner optimism may increase patient treatment outcome expectancies.
4. Increased perceptions of empathy, increased treatment outcome expectancies, and decreased anxiety, may increase patient enablement, satisfaction with the consultation, and health-related quality of life.
5. Increased perceptions of empathy, increased treatment outcome expectancies, and decreased anxiety, may decrease patient pain intensity, pain interference, and symptom severity.

**What (material):** This e-learning package includes text, images, and film. The materials are structured into the following sections:

**Introduction** (introduces the aims, benefits, and style of training, gives an overview of the content and duration of the e-learning, guidance on recording consultations, information on the team behind the training)
**Empathy** (presents latest research on aspects empathy including personalisation, validation, and increasing expressions of empathy as the consultation progresses; provides instruction and tips for expressing empathy to patients from unfamiliar cultures and in different situations, includes multimedia activities)
**Optimism** (presents latest research on outcome expectancies; provides phrases and techniques for conveying optimism at different points in the consultation, including positive safety netting; discusses what to do when optimism is difficult; includes multimedia activities)
**Osteoarthritis** (quiz to challenge empathy- and optimism-impairing myths in OA; provides specific examples of communicating clinical empathy and realistic optimism in consultations about OA; resources for practitioners and patients on OA)
**My Consultations** contains 3 guided activities: guidance on **how to reflect on recent consultations** and identify opportunities for improving the communication of clinical empathy and realistic optimism, with checklist; **goal setting** and **action planning**, for the user to set goals and make plans to change how they communicate clinical empathy and realistic optimism in consultations; **goal review** (presents user's personal goals back to them, prompts reflection on progress and invites revision/extension to goals).
**Resources** (direct access to video clips used in modules, checklist, OA resources, selected reading).

**What (procedures):** The e-learning package presents the Introduction section first, after which users can choose the order in which to review the Empathy, Optimism, and Osteoarthritis sections. After reviewing these sections, the user works through the My

Consultations sections covering reflection and goal setting and action planning; they can then access all prior sections from the menu. The goal review section is made available 4 weeks after the goal-setting section has been completed, and users are prompted about this via email. When working through the e-learning package, users are encouraged to engage in guided reflection, goal setting, and action planning.

**Who provided:** The intervention was provided remotely in the form of self-directed e-learning. As such, intervention recipients had no interaction with any 'providers' as such.

**How (mode of delivery; individual or group):** Intervention was delivered online via a website. Intervention recipients accessed it individually. There was no interaction with other recipients as part of the intervention.

**Where:** Intervention recipients accessed it using an internet connected device in a place of their choosing, e.g. at work, at home.

**When and how much:** Intervention recipients accessed it at a time of their choosing. The intervention was intended to take up to approximately 75 minutes to complete. It could be done all at once or in multiple sittings (user's choice).

**Tailoring:** No tailoring, although as described above recipients could choose the order in which they accessed some intervention sections.

**Modification:** No modifications were made to the intervention during this study.

**How well (planned):** There is no straight forward way to assess the fidelity of this self-directed e-learning intervention. Usage data were captured to give an indication of number of times and length of time people logged on to the intervention, and time spent logged on.

**How well (actual):** Practitioners accessed EMPathicO between one and eleven times (median = 2) and typically did so for between 30 and 120 minutes in total, spending longer on the content modules than on the reflection and goal setting modules.
